# Analysis of 3,760 hematologic malignancies reveals rare transcriptomic aberrations of driver genes

**DOI:** 10.1101/2023.08.08.23293420

**Authors:** Xueqi Cao, Sandra Huber, Ata Jadid Ahari, Franziska R. Traube, Marc Seifert, Christopher C. Oakes, Polina Secheyko, Sergey Vilov, Ines Scheller, Nils Wagner, Vicente A. Yépez, Piers Blombery, Torsten Haferlach, Matthias Heinig, Leonhard Wachutka, Stephan Hutter, Julien Gagneur

**Author notes:** Corresponding authors: Julien Gagneur Stephan Hutter Leonhard Wachutka.

## Abstract

**Background:** Rare oncogenic driver events, particularly affecting the expression or splicing of driver genes, are suspected to substantially contribute to the large heterogeneity of hematologic malignancies. However, their identification remains challenging.

**Methods:** To address this issue, we generated the largest dataset to date of matched whole genome sequencing and total RNA sequencing of hematologic malignancies from 3,760 patients spanning 24 disease entities. Taking advantage of our dataset size, we focused on discovering rare regulatory aberrations. Therefore, we called expression and splicing outliers using an extension of the workflow DROP (Detection of RNA Outliers Pipeline) and AbSplice, a variant effect predictor that identifies genetic variants causing aberrant splicing. We next trained a machine learning model integrating these results to prioritize new candidate disease-specific driver genes.

**Results:** We found a median of seven expression outlier genes, two splicing outlier genes, and two rare splice-affecting variants per sample. Each category showed significant enrichment for already well-characterized driver genes, with odds ratios exceeding three among genes called in more than five samples. On held-out data, our integrative modeling significantly outperformed modeling based solely on genomic data and revealed promising novel candidate driver genes. Remarkably, we found a truncated form of the low density lipoprotein receptor *LRP1B* transcript to be aberrantly overexpressed in about half of hairy cell leukemia variant (HCL-V) samples and, to a lesser extent, in closely related B-cell neoplasms. This observation, which was confirmed in an independent cohort, suggests *LRP1B* as a novel marker for a HCL-V subclass and a yet unreported functional role of *LRP1B* within these rare entities.

**Conclusions:** Altogether, our census of expression and splicing outliers for 24 hematologic malignancy entities and the companion computational workflow constitute unique resources to deepen our understanding of rare oncogenic events in hematologic cancers.

## Background

Hematologic malignancies are characterized by abnormal blood cells in the bone marrow, peripheral blood, or lymphatic organs. They can occur in various forms, affecting the myeloid or lymphoid cell lineage. In 2020, hematologic malignancies accounted for approximately 2.5% of new cancer cases globally and accounted for 3.1% of cancer-associated mortality (1). While some subtypes, like myeloproliferative neoplasm, exhibit a high degree of uniformity in their manifestation and genetic profile, others, like myelodysplastic neoplasm, display a significantly broader spectrum, hampering correct diagnosis and therapy decisions and negatively impacting treatment outcomes and survival (2). Thus, better understanding the variety of oncogenic events for each disease entity is of utmost interest to refine diagnostics and facilitate the development of new therapeutic options.

Within the last decade, the identification of driver genes in hematologic malignancies has been dramatically enhanced by the utilization of next-generation sequencing. This has provided valuable insights into the underlying genetic landscape of each entity and triggered a revision of the classification systems, which now emphasize genomics-based categorization of various leukemia and lymphoma entities (2–5). However, despite significant progress in understanding recurrent driver mutations in hematologic malignancies, much remains to be learned about the rare events within each disease entity that drive their individual development and progression (6,7).

Alterations of gene expression and splicing play a key role in cancer (8), particularly in hematologic malignancy pathogenesis (9–19). For instance, many hematologic malignancies are characterized by altered expression resulting from gene rearrangements that lead to the overexpression of specific transcription factors or cell cycle regulators. Examples include acute myeloid leukemia (AML) with defining genetic alterations (20), *BCR::ABL1*-positive chronic myeloid leukemia (CML) (21), or *CCND1* rearrangements in mantle cell lymphoma (MCL) (2,22). Moreover, aberrant splicing can generate gain or loss-of-function transcript isoforms of driver genes for many cancer types (23). For example, multiple aberrant splice isoforms of the *TP53* transcript have been observed in CML, even in the absence of genomic mutations around exon-intron junctions (24). However, a systematic analysis of rare expression and splicing aberrations among hematologic malignancies is still lacking.

To address this gap, we conducted a comprehensive analysis of genomes (Whole Genome Sequencing, WGS) and matched transcriptomes (total RNA sequencing, RNA-Seq) of tumor tissues from 3,760 patients spanning 24 hematologic malignancy entities (Figure 1). We analyzed this data using RNA-seq-based expression and splicing outlier callers, as well as AbSplice, a tool we recently published that predicts rare genetic variants causing aberrant splicing. We demonstrate how these results can be utilized to identify a novel marker for a rare entity and enhance the prediction of hematologic malignancy driver genes beyond the commonly used mutational recurrence. In summary, our study aims to deepen our understanding of the role of rare gene expression and RNA splicing in the development of hematologic malignancies and provide novel driver gene candidates.

**Figure 1.**
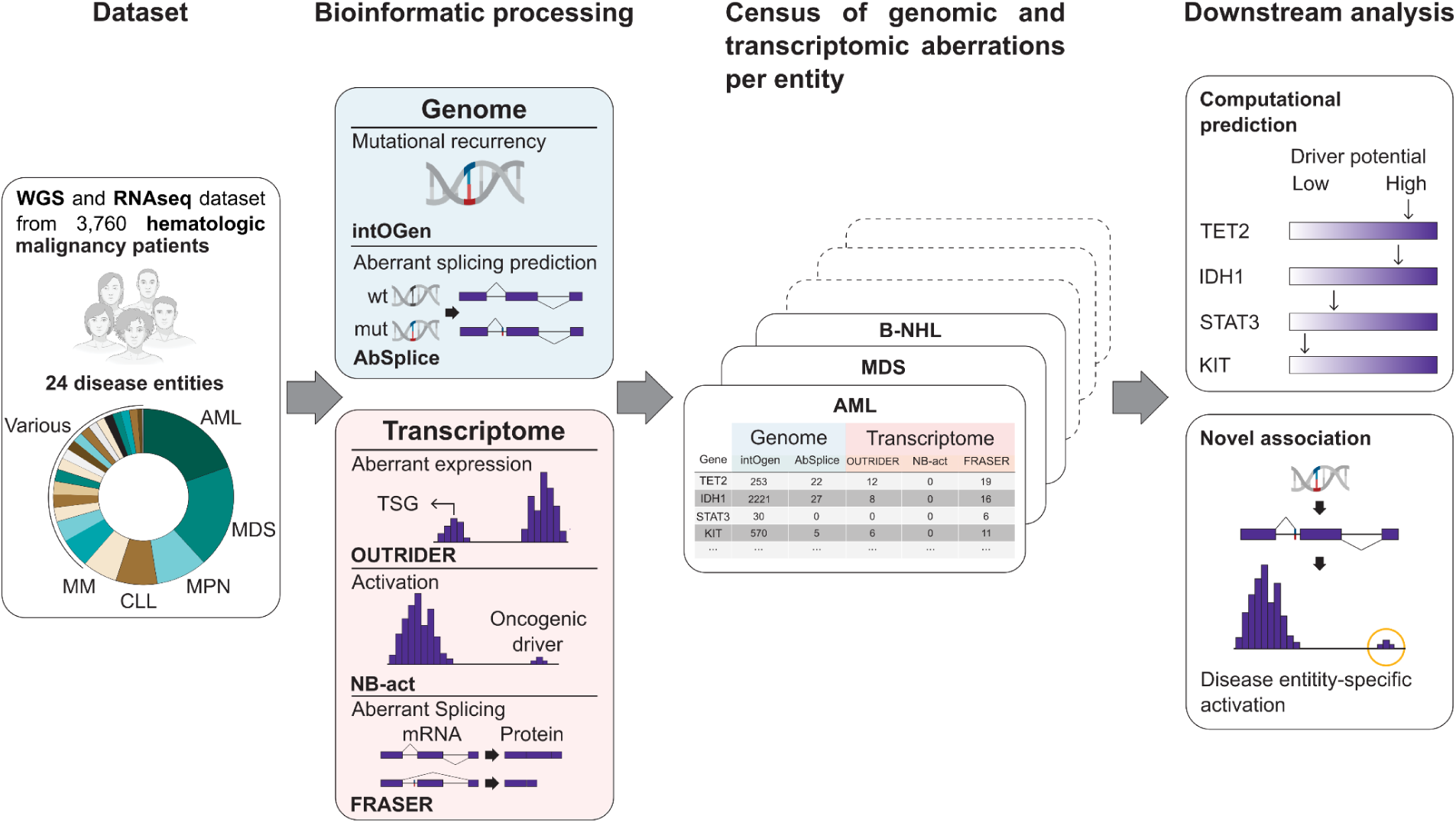
Overview of the study. Dataset: Whole genome sequencing and total RNA sequencing of 3,760 hematologic malignancies spanning 24 different disease entities. Bioinformatic processing: On genomic data, intOGen captures recurrent mutational patterns (37), and AbSplice predicts variants causing aberrant splicing (35). Working on RNA-seq data, OUTRIDER calls expression outliers of commonly expressed genes (39), NB-act calls overexpression of rarely expressed genes (Methods), and FRASER calls splicing outliers (41). Census: a unique collection of genomic and transcriptomic aberrations for 24 hematologic malignancy entities. Downstream analysis: driver gene prediction and enrichment analysis per disease entity.

## Methods

### Dataset

#### Patients

We used genomic and transcriptomic data from the Munich Leukemia Laboratory (MLL). We included a total of 3,760 tumor samples sent to the MLL between September 2005 and April 2019 for routine diagnostic workup (Table 1). Diagnoses from peripheral blood or bone marrow were based on cytomorphology, immunophenotype, cytogenetics, and molecular genetics, as previously described (25–27). All patients or their legal guardians gave written informed consent for genetic analyses and to the use of laboratory results and clinical data for research purposes, according to the Declaration of Helsinki. The study was approved by the MLL’s institutional review board. The dataset spanned 24 disease entities (Table 1).

**Table 1.**
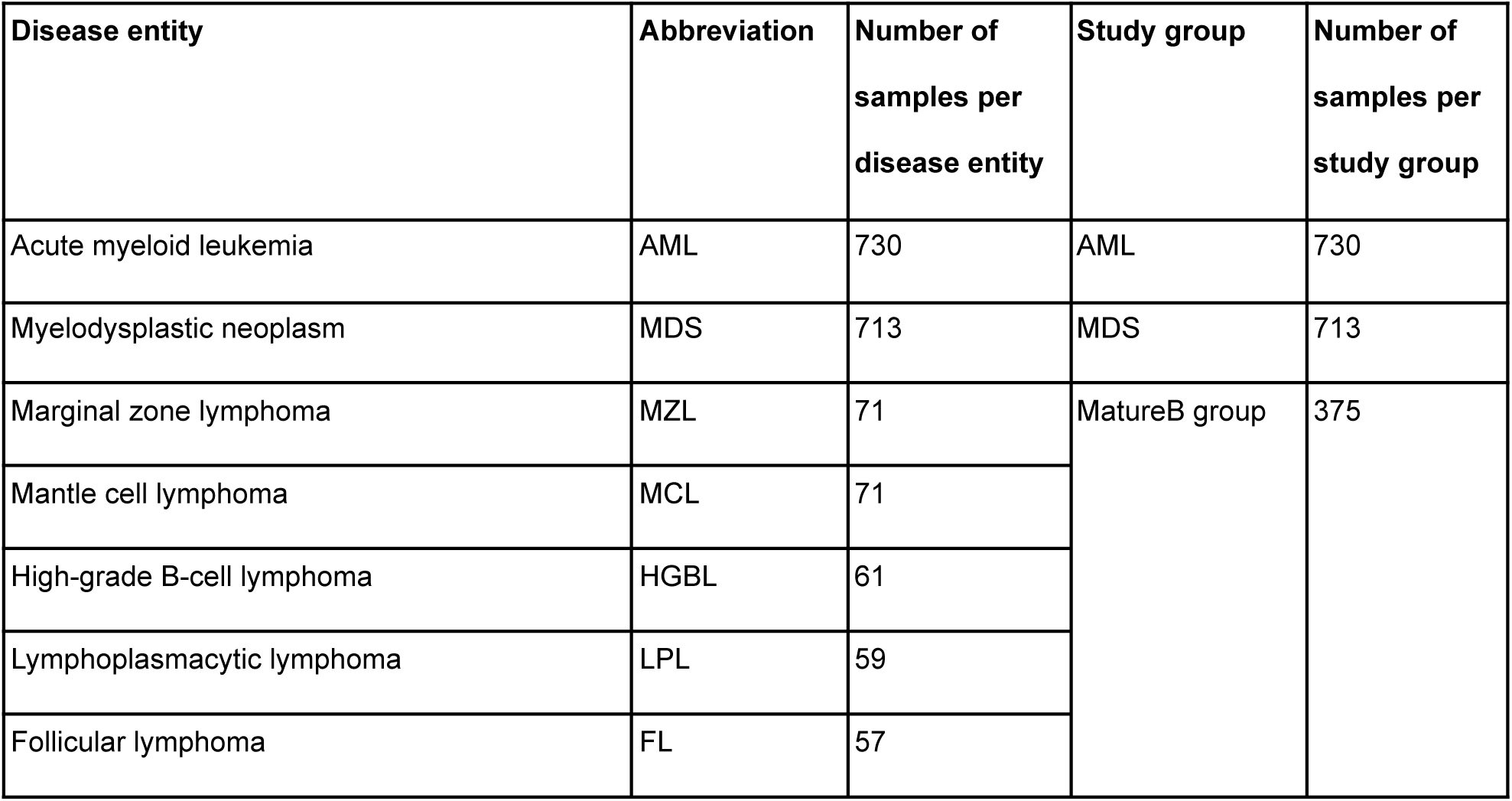
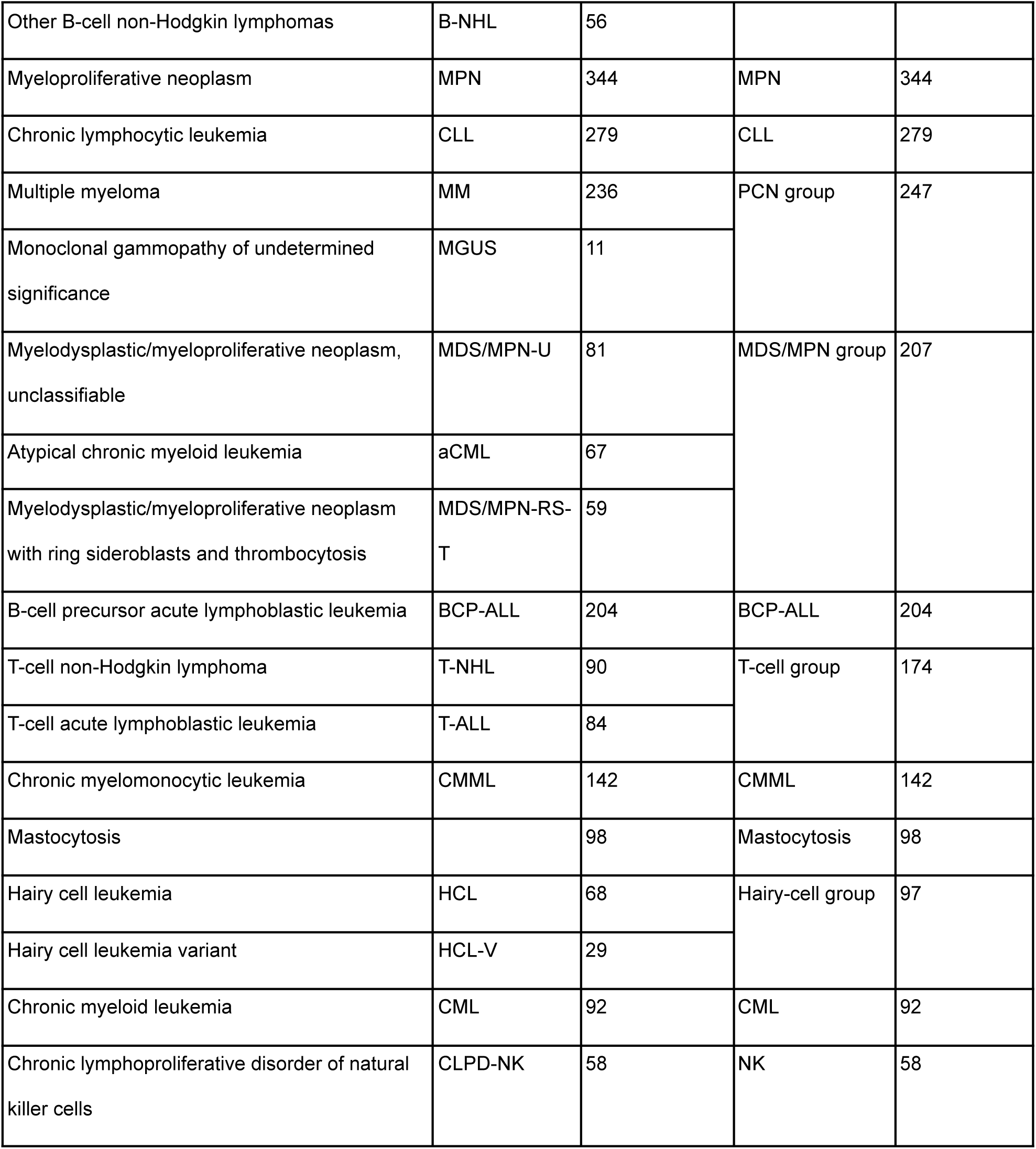
Disease entities enrolled and corresponding study groups.

| Disease entity | Abbreviation | Number of samples per disease entity | Study group | Number of samples per study group |
| --- | --- | --- | --- | --- |
| Acute myeloid leukemia | AML | 730 | AML | 730 |
| Myelodysplastic neoplasm | MDS | 713 | MDS | 713 |
| Marginal zone lymphoma | MZL | 71 | MatureB group | 375 |
| Mantle cell lymphoma | MCL | 71 |  |  |
| High-grade B-cell lymphoma | HGBL | 61 |  |  |
| Lymphoplasmacytic lymphoma | LPL | 59 |  |  |
| Follicular lymphoma | FL | 57 |  |  |
| Other B-cell non-Hodgkin lymphomas | B-NHL | 56 |  |  |
| Myeloproliferative neoplasm | MPN | 344 | MPN | 344 |
| Chronic lymphocytic leukemia | CLL | 279 | CLL | 279 |
| Multiple myeloma | MM | 236 | PCN group | 247 |
| Monoclonal gammopathy of undetermined significance | MGUS | 11 |  |  |
| Myelodysplastic/myeloproliferative neoplasm, unclassifiable | MDS/MPN-U | 81 | MDS/MPN group | 207 |
| Atypical chronic myeloid leukemia | aCML | 67 |  |  |
| Myelodysplastic/myeloproliferative neoplasm with ring sideroblasts and thrombocytosis | MDS/MPN-RS-T | 59 |  |  |
| B-cell precursor acute lymphoblastic leukemia | BCP-ALL | 204 | BCP-ALL | 204 |
| T-cell non-Hodgkin lymphoma | T-NHL | 90 | T-cell group | 174 |
| T-cell acute lymphoblastic leukemia | T-ALL | 84 |  |  |
| Chronic myelomonocytic leukemia | CMML | 142 | CMML | 142 |
| Mastocytosis |  | 98 | Mastocytosis | 98 |
| Hairy cell leukemia | HCL | 68 | Hairy-cell group | 97 |
| Hairy cell leukemia variant | HCL-V | 29 |  |  |
| Chronic myeloid leukemia | CML | 92 | CML | 92 |
| Chronic lymphoproliferative disorder of natural killer cells | CLPD-NK | 58 | NK | 58 |

#### Sample preparation

DNA and total RNA from peripheral blood and bone marrow samples were extracted using the MagNA Pure 96 Instrument and the MagNAPure96 DNA and Viral NA LV Kit and MagNA Pure 96 Cellular RNA LV Kit, respectively (Roche LifeScience, Mannheim, Germany). WGS and RNA-seq were performed on the prepared samples (Supplementary Materials and Methods).

#### Variant calling and annotation

Variant calling of single nucleotide variants, short insertions and deletions, structural variants, copy number variations, and gene fusions were performed on all samples as described previously (28–31) (Supplementary Materials and Methods). The analysis was based on the GENCODE v33 (32) annotation and using the reference genome GRCh37.

As matched germline tissues for the tumor samples were unavailable, sex-matched genomic DNA from random healthy donors was used as normal material during tumor-normal variant calling. To reduce germline variants and artifacts from the data, we filtered single nucleotide variants, short insertions, and deletions with the following criteria:

1. Only consider variants emitted with “PASS” quality
2. Discard variants with gnomAD v2.1.1 (33) minor allele frequency > 0.05%
3. Discard variants with sample variant allele frequency < 0.1

To reduce germline variants and artifacts from structural variant calls we only considered variants meeting the following criteria:

1. Only consider variants emitted with “PASS” quality
2. Only consider variants if they have three or more paired-reads supporting it
3. Discard variants found with exact breakpoint locations in the gnomAD SV database (34)
4. Discard variants that are found in four or more different myeloid and four or more different lymphatic entities

To predict the effects of variants on splicing, we applied AbSplice-DNA v1.0.0 (35) to the filtered variants. AbSplice-DNA estimates the probability that a rare variant causes aberrant splicing in a given tissue, which takes the variant and splice annotations, so-called SpliceMaps, as input.

We generated SpliceMaps for each study group as described previously (35). The variants with an AbSplice-DNA prediction score ≥ 0.2 were classified as splice-affecting variants. Further variant effect predictions were obtained by applying Ensembl VEP v81 (36). VEP splice-related variants were defined as VEP calculated consequences ‘splice_acceptor_variant’ (2 base region at the 3’ end of an intron), ‘splice_donor_variant’ (2 base region at the 5’ end of an intron), and ‘splice_region_variant’ (1-3 bases of the exon or 3-8 bases of the intron).

Gene-level mutational recurrence scores were obtained using IntOGen commit 437a047 (37), and MutSigCV v1.41 (38). To this end, the variants were additionally filtered using the following criteria:

1. Discard variants with sample variant allele frequency < 0.15 as this more stringent cutoff led to improved driver gene enrichments
2. Only consider variants with VEP calculated impact ‘HIGH’, ‘MODERATE’, ‘LOW’ or calculated consequence ‘5_prime_UTR_variant’, ‘3_prime_UTR_variant’, and ‘coding_sequence_variant’
3. Only consider variants supported by at least 20 reads

#### Expression and splicing outlier calling

We implemented a python backend version of OUTRIDER v.1.99.0 (39) for scalability (see Availability of data and materials) and applied it to RNA-seq data using DROP v1.1.0 (40) w(40)default settings to call expression outliers at a False Discovery Rate (FDR) of 0.05. Expression outliers with lower-than-average expressions (z-score < 0) were defined as underexpression outliers, and higher-than-average expressions (z-score > 0) as overexpression outliers.

FRASER v.1.99.0 (41) was applied to RNA-seq data using DROP v1.2.3 (40) with default settings to call splicing outliers (FDR < 0.05). We grouped the samples into 14 study groups based on hematopoietic cell origins and pathologies (2–5) to ensure sufficient sample sizes (Table 1, N at least 58). Splicing outliers with delta Intron Jaccard Index >0 were defined as overrepresented splicing outliers, and delta Intron Jaccard Index <0 as underrepresented splicing outliers.

As OUTRIDER is restricted to commonly-expressed genes defined as genes with fragments per kilobase of transcript per million mapped reads (FPKM) larger than 1 in at least 5% of the samples, we introduced a novel method to detect aberrant activation of genes usually not expressed. This method, NB-act (Negative Binomial activation), provides *P*-values for observed fragment count (read pairs) for each gene in each sample under the null hypothesis that the gene is not expressed in the sample. Specifically, NB-act computes the probability of observing a certain number of fragments or more for a gene in a particular sample, assuming a negative binomial distribution with an expected baseline expression of 1 FPKM and a dispersion parameter of 0.02 (Supplementary Materials and Methods). The FPKM value of 1 corresponds to the threshold separating expressed from non-expressed genes in OUTRIDER (39). The dispersion parameter of 0.02 corresponds to the empirically observed lowest dispersion values estimated by OUTRIDER on expressed genes. As low dispersion corresponds to high variance, we chose a low dispersion value for NB-act to be conservative. NB-act was applied to rarely-expressed genes (FPKM>1 in at least one sample but less than 5% of the samples).

#### Enrichment for driver gene and variant categories

The enrichment of cancer driver genes was evaluated by the Fisher test using Cancer Gene Census (CGC) GRCh37 v97 (42) (Supplementary Materials and Methods). The role of hematologic malignancy driver genes was determined using annotation ‘Tissue Type’ and ‘Role in Cancer’.

As promoter variants, we considered all single nucleotide variants and short insertions and deletions less than 2,000 bp away from the transcription start site of the corresponding gene. Frameshift and stop gained variants were detected with Ensembl VEP v81. The enrichment analysis of variants within expression outlier gene-sample pairs was performed using the Fisher test. Sample-gene pairs with other variants were excluded apart from those for which enrichment was calculated.

### Survival analysis

Overall survival analyses were performed according to Kaplan-Meier and compared using two-sided log-rank tests with the statistical software R v4.2.2 with the survival (43) and survminer (44) packages. The overall survival was calculated as the time from diagnosis to death or last follow-up.

### Driver gene prediction

Machine-learning models were trained to predict the probability of a gene to be a hematologic malignancy driver gene. To this end, random forest classifiers (Python package scikit-learn v1.0.2) (45) were trained to predict driver genes using gene-level features obtained from all samples on the one hand and each of the 14 study groups on the other hand. The gene-level features consisted of 21 features from seven gene-level metrics of seven intOGen tools, nine features from AbSplice-DNA scores, 22 features from OUTRIDER obtained using combinations of fold-change direction, significance, and effect size cutoffs, and, similarly, 11 features from NB-act and 22 features from FRASER (Supplementary Materials and Methods). In total, 377 genes listed among the hematologic panel genes (Supplementary Materials and Methods) or the hematologic malignancy driver genes from CGC GRCh37 v97 were used as the positive class for the classifiers. The random forests were trained using the function ‘RandomForestClassifier’ with the minimum of number of samples required to split an internal nodes set to 19, the maximal tree depth set to 10, and default settings otherwise (Supplementary Materials and Methods). The models were trained with 5-fold cross-validation, stratified to preserve the percentage of positive class in all folds, using the function ‘StratifiedKFold’ from the Python package scikit-learn v1.0.2. Performances were evaluated by precision-recall curves on the held-out data. Subsequently, only prediction values on held-out data were used for candidate curation. As the predicted probabilities range very differently across study groups, no fixed threshold was set, and we focused on the rank of the predicted genes. The benchmark against intOGen and MutSigCV was conducted on all study groups.

Further detailed descriptions are available in the Supplementary Materials and Methods.

### Validation HCL-V dataset

#### Patients

The validation HCL-V dataset included a total of 42 patients, including 14 HCL-V and 28 HCL patients. The HCL-V patients were diagnosed based on immunophenotype and morphological characteristics consistent with HCL-V, including all CD5 negative, CD11c positive, and CD123 negative markers. All HCL-V patients were confirmed as *BRAF*-V600E negative.

#### Sample preparation

Tumor cells were isolated by cell sorting (purity > 99%) as FSC^high^CD20^+^CD11c^+^ and kappa/lambda light chain restriction. RNA was isolated by the RNeasy Micro Kit (Qiagen, Hilden, Germany).

#### RNA sequencing, read mapping

For the generation of RNA-sequencing libraries, the NuGEN Trio RNA-Seq System (NuGEN, Redwood City, California) was used. Samples were split equally and processed in independent sequencing steps to allow for the correction of batch effects. Sequencing was performed with paired-end sequencing and two times 100-bp length. Sequences were aligned with HiSAT2 v2.1.0 (46) to the GRCh38.

#### Gene expression analysis

Gene expression was analyzed with DESeq2 (47). The size factor was calculated using DEseq2. The counts were normalized by size factors. The activation of *LRP1B* was determined with a two-component Gaussian mixture clustering with equal variance on size factor-normalized and log-transformed counts, using the R package ‘mclust’ v.6.0.0 (48).

## Results

We investigated WGS and RNA-seq data from 3,760 tumor samples representing 24 different types of leukemia and lymphoma (Table 1). This is the largest collection of hematologic malignancy samples with WGS and matched RNA-seq, which also includes rare disease entities like hairy cell leukemia variant (HCL-V) and chronic lymphoproliferative disorder of natural killer cells. In order to restrict our analysis to putative rare germline and somatic variants, we filtered variants called on WGS with stringent quality filters and population allele frequency (33). We next annotated the genes using the seven features from the software intOGen, which include positional recurrence of mutations in genome sequence (OncodriveCLUSTL), positional recurrence of mutations in protein conformation (HotMAPS), enrichment of mutations in functional domains (smRegions), three alternative measures of selection strength inferred from synonymous and nonsynonymous mutations (CBaSE, MutPanning, and dNdScv), and OncodriveFML, a method identifying excess of somatic mutations across tumors in both coding and non-coding genomic regions (37,49–55). Moreover, we annotated genetic variants falling into gene bodies, including deep intronic variants, with AbSplice-DNA, a tool predicting variants causing aberrant splicing (35). On the RNA-seq data, we used OUTRIDER on a total of 12,966 protein-coding genes commonly expressed across the dataset to call high or low expression outliers, and FRASER to call splicing outliers (39,41). We also introduced a new method, NB-act, to call rare aberrant activation of genes mainly not expressed (Methods). Combining all these results, we established a unique census of genomic and transcriptomic aberrations in 3,760 hematologic malignancy samples (Figure 1).

### Expression outliers are enriched for hematologic malignancy driver genes

Case-control differential expression analysis identifies common differences between tumor and healthy samples and, therefore, can be suited to identify recurrently differentially expressed genes in tumors. To identify potential rare driver genes, we instead employed expression outlier analysis, which calls rare, aberrantly high, or low expression of a gene within a dataset. OUTRIDER calls expression outliers by modeling read count distribution across samples and reporting the statistical significance of extreme observations. Moreover, OUTRIDER controls for gene expression covariations, which allows automatic corrections for technical sources of variation, such as batch effects, and adjusts for transcriptome-wide co-regulation patterns due to trans-acting regulatory changes. Deviations from these variations have been shown to be enriched for rare variants with strong cis-regulatory effects (39) and help identify causes of rare disorders (56–59). Applying OUTRIDER, we called 21,264 underexpression outliers (median of 2 per sample) and 14,041 overexpression outliers (median of 2 per sample) on 10,193 different protein-coding genes (Supplementary Figures 1-2). To verify whether these outliers are associated with cancer, we calculated their enrichment for reported hematologic malignancy driver genes, which we adapted from CGC (42). We observed a strong enrichment for CGC hematologic tumor suppressor genes among underexpression outliers and for CGC hematologic oncogenes among overexpression outliers (Figure 2A-B). Notably, the genes called as outliers in more than five samples exhibited the highest enrichment, indicating that genes frequently called as outliers are more likely to be oncogenic. Moreover, we found that the number of expression outliers per sample was unevenly distributed (Supplementary Figures 1-2). Samples with numerous outliers may either represent cases where OUTRIDER could not adequately fit the data or situations where the gene regulatory network is globally affected, resulting in widespread expression aberrations throughout the genome. We reasoned that the enrichment for driver genes among expression outliers could be lower in all those samples. Indeed, the enrichment for tumor suppressor genes and oncogenes increased when focusing on the three most significant outliers (at most three) in each sample (Figure 2A-B).

**Figure 2.**
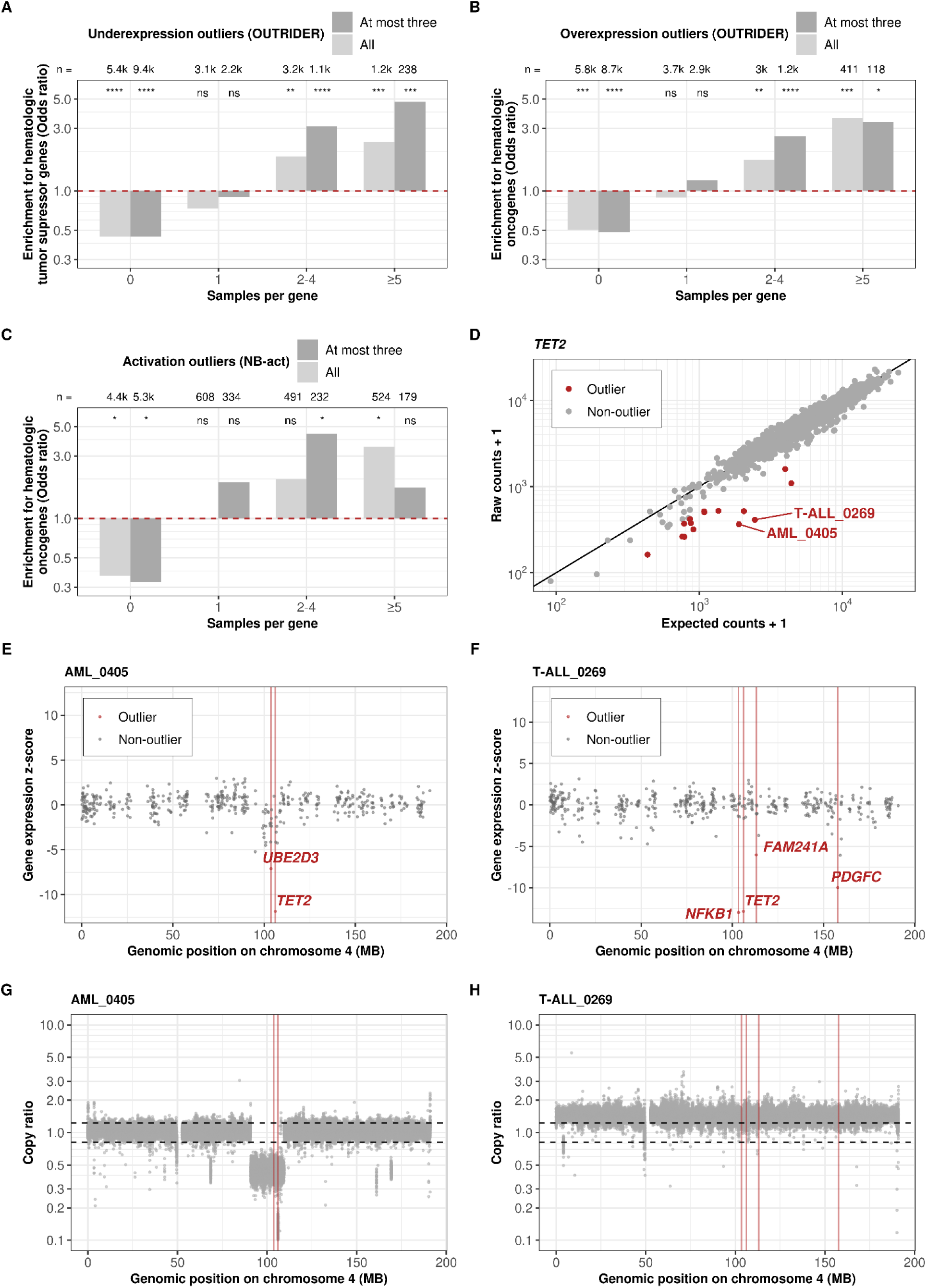
Expression outliers are enriched for hematologic malignancy driver genes. **(A)** Enrichment for CGC hematologic tumor suppressor genes among all genes called by OUTRIDER as well as at most three significant genes per sample that were called to be underexpression outliers. The genes are stratified by the number of samples in which the gene is called as an outlier. Numbers of the genes and nominal significances from the Fisher test are labeled at the top of the bars (ns: not significant; *: *P* ≤ 0.05; **: *P* ≤ 0.01; ***: *P* ≤ 0.001; ****: *P* ≤ 0.0001). **(B-C)** As in A) for CGC hematologic oncogenes among overexpression and activation outliers, respectively. **(D)** Raw RNA-seq read counts per sample against expected counts for *TET2*. For fifteen samples (red), *TET2* was called as an underexpression outlier, including samples AML_0405 and T-ALL_0269, showcased in the panels (E-H). **(E-F)** Gene expression z-score among all samples against the genomic position of the genes on chromosome 4. **(G-H)** Copy ratio within chromosome 4 binned into one-kb region. Red vertical lines mark the genomic position of the expression outliers. Black dashed lines mark the estimated region of no copy number variation. The underexpression outliers in *TET2* and *UBE2D3* in sample AML_0405 reflect the consequence of copy number loss that is very specific to the *TET2* locus. In contrast, an extra copy of whole chromosome 4 (karyotype: 47,XY,+4) is found in sample T-ALL_0269. *TET2* reduced expression must have a different cause in this sample.

OUTRIDER filters out genes expressed in less than 5% of samples due to statistical modeling limitations, leaving a gap in detecting rare gene activation. To fill this gap, we developed a complementary algorithm, NB-act (Methods). We applied it to the 6,017 rarely-expressed protein-coding genes filtered out by OUTRIDER. NB-act identified 10,263 activation outliers among 1,623 genes (with a median of 0 and 75% quantile of 2 per sample, Supplementary Figure 3). We observed a notable enrichment for CGC hematologic oncogenes among all activation outliers (Figure 2C). Here too, restricting to at most three outliers per sample increased the enrichment. Altogether, these analyses provide a unique set of aberrantly expressed genes in hematologic malignancies with strong enrichment for driver genes.

We next investigated how expression outliers were associated with genomic alterations. We observed a strong enrichment of relevant genomic aberrations among the expression outliers. Overall, 18.8% of the underexpression outliers could be explained by a copy number loss, and 11.4 % of the overexpression outliers and 10.2% of the activation outliers could be explained by a copy number gain (Supplementary Figure 4). We also found significant enrichments for rare variants associated with nonsense-mediated decay (stop-gained, frameshift, and splice-related) as well as structural variants and variants found in the promoter region among underexpression outliers (Methods, Supplementary Figure 5), consistent with earlier reports in non-cancer samples (60). Structural variants were also significantly enriched among overexpression and activation outliers (Supplementary Figure 6-7). Interestingly, we found enrichments for rare splice-related variants and rare frameshift variants among overexpression outliers but not activation outliers, perhaps because, in some cases, these variants lead to RNAs with increased stability (Supplementary Figure 6-7). Overall, these enrichments for relevant genomic aberrations support the reliability of the expression outlier calls and provide a genetic explanation for a substantial fraction of them.

Investigations of expression outlier events of the tumor suppressor gene *TET2* illustrate how this catalog can be used. Loss of function of *TET2* has been reported in myeloid leukemia due to splice site mutations, out-of-frame insertions or deletions, and base substitutions (61–64). We found 15 underexpression outlier events (two in AML, one in myelodysplastic neoplasm, seven in B-cell precursor acute lymphoblastic leukemia, and five in T-cell acute lymphoblastic leukemia) for *TET2* across all samples and no overexpression outliers (Figure 2D), consistent with its role as a tumor suppressor gene. To gain further insights, we showcased the two samples with the lowest fold changes (samples AML_0405 and T-ALL_0269). For these two samples, *TET2* was among the most extreme outliers (Supplementary Figure 8-9). However, neither the single nucleotide variants, short insertions and deletions, structural variants, nor gene fusions explained the observed decrease in *TET2* expression. Inspection of the genomic coverage indicated a loss of the *TET2* locus nested within a single-copy loss of a larger region in chromosome 4 for sample AML_0405. This explanation was further supported by the decreased expression of the neighbor gene *UBE2D3* (Figure 2 E, G). In contrast, genomic coverage investigations did not provide an explanation for the reduced expression of *TET2* in sample T-ALL_0269. Further outliers, including *TET2* in sample T-ALL_0269, could reflect yet-to-be-interpreted genetic variants, epigenetic causes, or outlier caller false positives (65).

In summary, both OUTRIDER and NB-act reveal aberrantly expressed genes enriched for driver genes and can be used to identify downregulated tumor suppressor genes or activated oncogenes in individual samples.

### Rare splicing aberrations are enriched for hematologic malignancy driver genes

We applied FRASER to detect aberrant splicing events (aberrant usage of existing or novel splice sites) within our samples, which could be caused by events such as alternative exon usage, intron retention, alternative donor or acceptor site usage, usage of deep intronic donor and acceptor sites or truncation of parts of the transcript. Like OUTRIDER for expression, FRASER is a tool to call splicing outliers while controlling for covariations, a task that is distinct from calling differential splicing between groups (41). We called 43,464 splicing outliers across 35,410 gene-level splicing outlier events on a total of 7,591 genes in 2,854 samples (Supplementary Figure 10). Remarkably, we observed a substantial enrichment for CGC hematologic tumor suppressor genes among these splicing outliers (Figure 3A). As for expression outliers, restricting to at most three outliers per sample increased the enrichment. Moreover, the genes called as splicing outliers in more than five samples exhibited the highest enrichment, suggesting a higher potential of being oncogenic.

**Figure 3.**
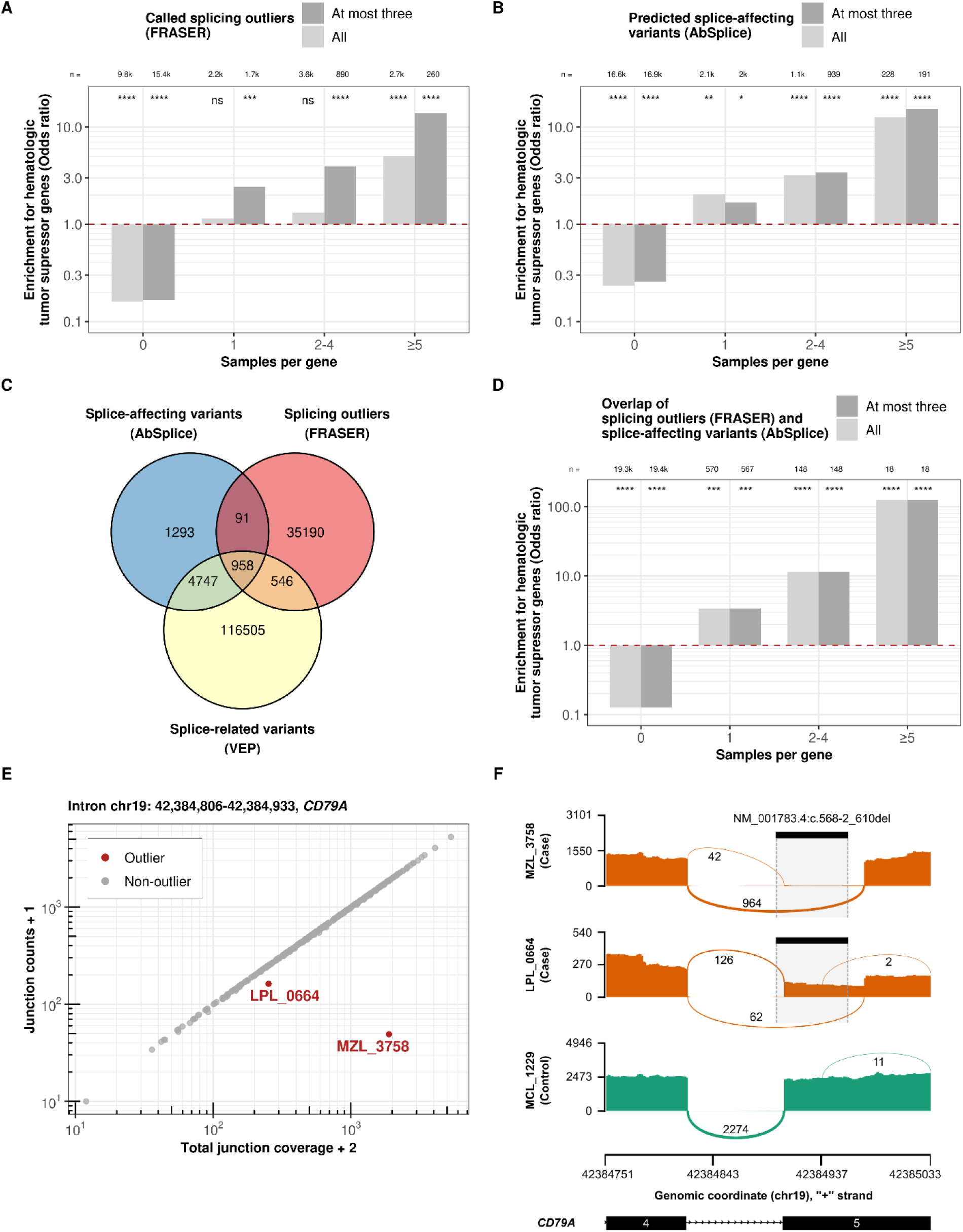
Transcriptomic splicing outliers and genomic splice-affecting variants are enriched for hematologic malignancy driver genes. **(A)** Enrichment for CGC hematologic tumor suppressor genes among all genes called by FRASER and at most three significant genes per sample that were called aberrantly spliced from RNA-seq (FRASER). The genes are stratified by the number of samples in which a gene is called as a splicing outlier. Numbers of the genes and their nominal significance from the Fisher test are labeled at the top of the bars. **(B)** As in (A) among splice-affecting variants (AbSplice). **(C)** Overlap of splice-affecting variants, VEP splice-related variants, and splicing outliers on the sample-gene level. **(D)** As in (A) among splice-affecting variants with corresponding splicing outliers. **(E)** Junction counts (split reads) against total junction coverage (exon-intron or intron-exon spanning reads) of the displayed intron. The displayed intron of the *CD79A* only shows aberrant splicing in samples MZL_3758 and LPL_0664. **(F)** Sashimi plots showing RNA-seq read coverage (y-axis) and the numbers of split reads spanning an intron indicated on the exon-connecting line (using pysashimi) for two aberrant splicing events. Two case samples using a unique splice-site in *CD79A* 5^th^ exon acceptor site and one control sample are displayed. The rare splice-affecting deletion (NM_001783.4:c.568-2_610del) predicted by AbSplice, which existed exclusively in the two case samples, is shown as black bars.

Calling splicing aberrations from RNA-seq data can point toward mutations that have been overlooked using the genomic data alone. As an example, we showcase RB1, a well-known tumor suppressor gene, which can be inactivated by various mechanisms, including intragenic mutations, methylation of the promoter region, and chromosomal deletions (66). In sample B-NHL_1747, we identified an unusual exon-skipping event of the 20^th^ exon in RB1 (Supplementary Figure 11). Analyzing the WGS data of this sample revealed a deletion of 118-base-pair in the region of the 20^th^ exon, which led to a frame-shift exon-skipping (Supplementary Figure 12). However, this structural variant was initially discarded in our variant calling pipeline due to stringent filtering. Using FRASER, which accurately captured the consequence of this initially discarded structural variant as a splicing outlier, we were now able to recognize its significant transcriptomic impact and rescue it.

As a complementary approach to RNA-seq-based splicing outlier calling, we considered AbSplice (35), a recently published algorithm that predicts whether a rare variant causes aberrant splicing. Here, we applied it for the first time to hematologic malignancy samples. AbSplice integrates two sequence-based machine learning tools, MMSplice (67) and SpliceAI (68), with so-called SpliceMaps, which are quantified tissue-specific usages of splice sites, including non-annotated and weak splice sites. We derived SpliceMaps from the raw RNA-seq data (Methods). AbSplice classified 7,160 rare variants as splice-affecting out of 275,899,978 pre-filtered variants, resulting in 7,074 genes predicted to be affected across 3,093 samples (with a median of 2 per sample, Supplementary Figure 13). These results demonstrated a strong enrichment for CGC hematologic tumor suppressor genes among all genes carrying splice-affecting variants (Figure 3B). Furthermore, when focusing on the genes that were predicted to have recurrent aberrant splicing events across multiple samples, we observed an even higher enrichment, indicating a tendency for recurrent splice-affecting variants to occur in known hematologic malignancy driver genes (Figure 3B). In contrast to the RNA-seq-based splicing outlier calls, restricting AbSplice predictions to at most three calls per sample did not lead to a significantly higher enrichment, likely due to the limited number of samples with a high number of variants displaying strong AbSplice scores. Collectively, these results indicate that the rare splicing aberrations predicted from the genome can contribute to identifying potential driver genes in hematologic malignancies.

Overall, 15% of the AbSplice predictions (1,049 out of 7,089, Figure 3C) were also called as splicing outliers by FRASER, which is lower but not far off the claimed precision of AbSplice at the cutoff we used (predicted precision cutoff = 0.2). This proportion of FRASER splicing outliers was much larger using AbSplice than when using VEP splice-related rare variants (1.2%, 1,504 out of 121,256, Figure 3C). Among those 1,049 gene-sample pairs common to AbSplice and FRASER, the enrichment for CGC hematologic tumor suppressor genes was substantially stronger than when considering AbSplice predictions or FRASER calls separately (Figure 3D). As an example of an event both predicted by AbSplice and called by FRASER, we showcase *CD79A*, a well-known oncogene frequently affected by somatic mutations in hematologic malignancies (42,69–71). We discovered splicing outliers of *CD79A* in samples MZL_3758 and LPL_0664, resulting in a truncation of 18 amino acids at the beginning of the 5^th^ exon (Figure 3E). AbSplice predicted that a rare 45-base-pair deletion spanning the acceptor site (NM_001783.4:c.568-2_610del) to be splice-affecting (AbSplice score = 0.36, Figure 3F, Supplementary Figure 14). This deletion, which was exclusively found in these two samples, is very likely the cause of this aberrant splicing event. Notably, similar deletions in the 4^th^ and 5^th^ exon of *CD79A* have been observed in diffuse large B-cell lymphoma, which has been shown to impact the ITAM signaling modules (69).

In summary, variants predicted to cause aberrant splicing by AbSplice and splicing outliers called by FRASER in transcriptomic data showed global enrichment for driver genes and proved invaluable in pinpointing affected driver genes in individual samples.

All our results from splice-affecting variants, expression outliers, splicing outliers, and recurrent mutational patterns aggregated by disease entities are provided (Availability of data and materials). This constitutes a comprehensive census detailing genomic and transcriptomic aberrations across 3,760 hematologic malignancy samples.

### Integration of rare genomic and transcriptomic aberrations improves hematologic malignancy driver gene prediction

We next trained models to predict driver genes based on genomic and transcriptomic features, including the seven intOGen tools, AbSplice, OUTRIDER, NB-act, and FRASER. To this end, we used the complete dataset on the one hand and used the dataset stratified by 14 study groups on the other hand (Table 1). As ground truth, we considered the 322 CGC hematologic malignancy driver genes complemented with 55 additional curated hematologic panel genes (Methods). The performance of all 15 models was evaluated using 5-fold cross-validation (Methods).

Using the complete dataset, we found that the genomic and transcriptomic features exhibited complementary predictive value for hematologic malignancy driver genes (Figure 4A-B, Supplementary Figure 15-16). Specifically, integrating AbSplice variant effect predictions significantly enhanced the genomic-based model trained on the seven intOGen tools. Moreover, we found that the transcriptomic features further significantly improved the model (Figure 4B). These findings underscore the relevance of incorporating aberrant expression and splicing analyses to predict driver genes. Among the 100 top-ranked genes, 63 were known from the ground truth, vastly greater than expectation (odds ratio = 106.3, *P* = 1.6×10^-84^, Fisher test; Figure 4C, Supplementary Table 1). These enrichments for well-known driver genes demonstrated the reliability of the model predictions. Four genes, *CDNKN1B*, *EIF3E*, *HLA-A,* and *IL6ST*, are CGC driver genes that have not been annotated as hematologic yet, indicating a broader role for those. Despite *TTN* being known to be false positive in mutational recurrence analysis as a long gene, the remaining genes were categorized as candidate drivers whose role remains to be further assessed.

**Figure 4.**
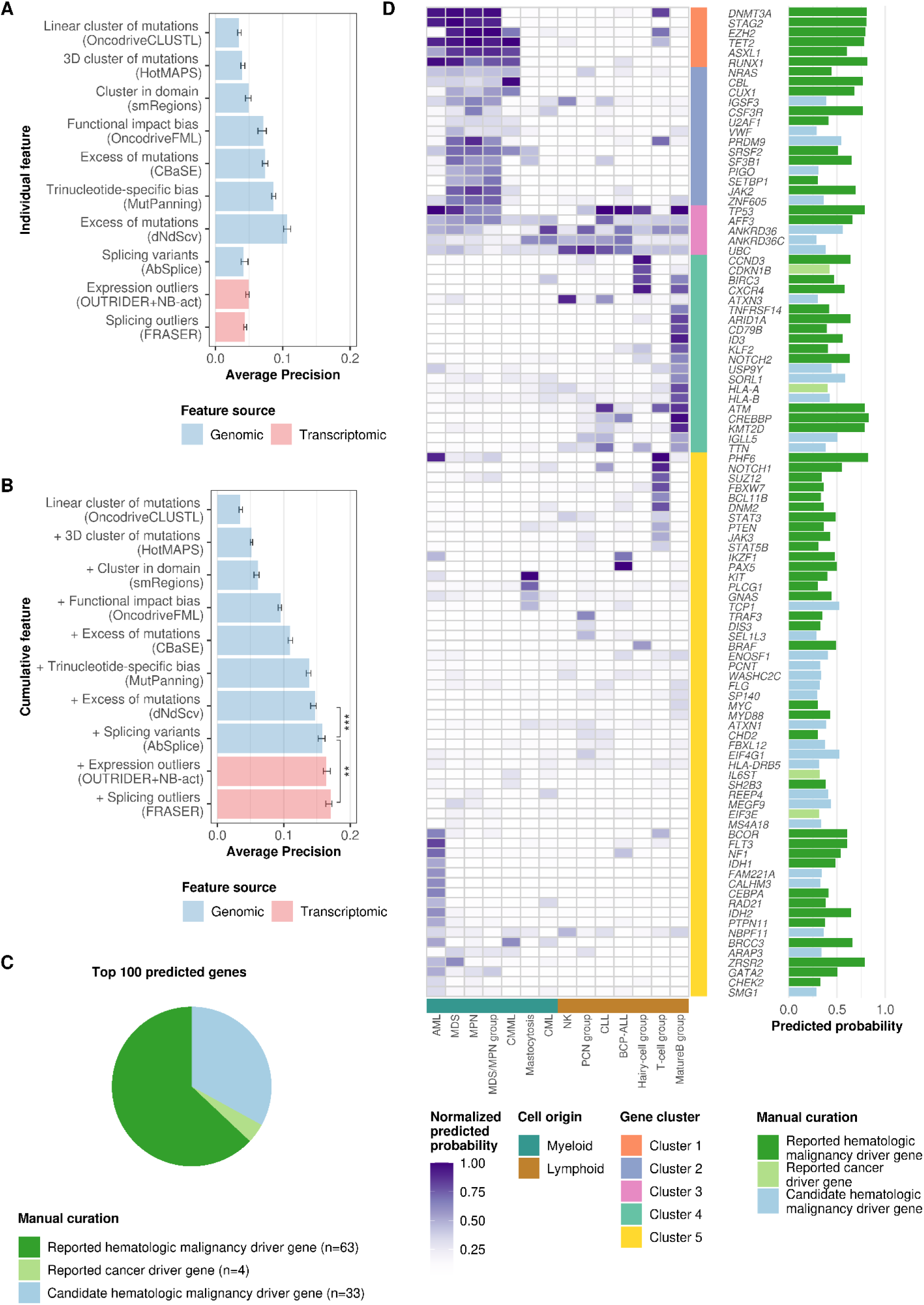
Rare genomic and transcriptomic aberrations add value to hematologic malignancy driver gene prediction on top of methods detecting mutational recurrence. **(A)** Individual features and the corresponding model’s performances (measured by average precision) using a random forest classifier. **(B)** Order as in (A) for cumulative features. The performance constantly improved when adding additional features. Asterisks denote nominal significance from the Wilcoxon test. **(C)** Numbers of genes in each category among the top 100 predicted genes when using all features and the complete dataset. Reported hematologic malignancy driver genes are the genes listed in either CGC hematologic malignancy driver genes or hematologic panel genes. Reported cancer driver genes are the genes documented by CGC. The rest of the genes are categorized as candidate hematologic malignancy driver genes. **(D)** The heatmap shows the predicted driver gene probability per gene (rows) and study group (columns) relative to the column-wise maximum value. Myeloid and lymphoid entities clustered due to the shared hematologic malignancy driver gene profiles (bottom track). The barplot shows the predicted probabilities from the model trained on the complete dataset. Bar colors as in (C).

In line with the results on the complete dataset, our integrative model outperformed or was on par with intOGen or MutSigCV - a driver gene predictor based on mutation frequency across all study groups (Supplementary Figure 17). The study-group-wise models allowed further insights into disease-entity specificities (Figure 4D, Supplementary Table 2). Clustering the 100 top-ranked genes according to their study-group predicted probabilities revealed various disease entity specificities. Cluster 3 consisted of genes that exhibited high predicted probabilities across all disease entities. While *TP53* is a well-known pan-cancer tumor suppressor, we acknowledge that *ANKRD36*, *ANKRD36C*, and *UBC* may warrant further scrutiny as potential artifacts in global analysis (72). Clusters 1 and 2 comprised genes that scored highly specifically in myeloid disease entities, such as *ASXL1* and *SRSF2* (4,73). Conversely, the genes in the remaining clusters were associated with specific disease entities. Notable examples include *NOTCH1, PHF6, and FBXW7,* which are well-acknowledged for their role in T-cell leukemia, as well as *BRAF*, which was exclusively predicted in the hairy-cell group, in accordance with its recognized function as a marker for HCL (4,74–76).

Among the candidate genes, we identified several promising genes whose roles in hematologic malignancies are yet to be established. Our analysis predicted *SORL1* as a candidate driver in multiple study groups, including myelodysplastic neoplasm (precursor of AML), B-cell precursor ALL, and a study group comprising T-cell non-Hodgkin lymphoma and T-cell acute lymphoblastic leukemia. Consistent with these observations, *SORL1* was found to be expressed on the leukemic cell surface and released into plasma in AML and ALL, with its level decreasing during remission (77). *EIF4G1* stood out as another interesting candidate driver in AML and lymphoid entities, supported by previous analyses suggesting that *EIF4G1* is involved in cell survival in AML as a downstream target of *MYCN*, a known oncogene in neuroblastoma (78). We also found *TCP1* to be predicted in mastocytosis and B-cell precursor-ALL, whose high expression has been associated with AML drug resistance and poor survival through the activation of *AKT/mTOR* signaling (79). A fourth exciting candidate, *ATXN1,* was predicted as a candidate driver in multiple disease entities. Its important paralog *ATXN1L* is known as a novel regulator of hematopoietic stem cell quiescence (80). Overall, our predictions based on 3,760 tumor genomes and transcriptomes reveal promising hematologic malignancy driver candidates.

### Outlier clustering identifies *LRP1B* as a potential marker in HCL-V and related B-cell malignancies

In addition to our global driver gene prediction analysis, we performed a detailed investigation into each disease entity, examining its association with expression outliers, splicing outliers, and variants predicted to cause aberrant splicing. Overall, we found 2,716 significant associations between 11,273 genes and 24 disease entities (Supplementary Table 3, Benjamini-Hochberg false discovery rate < 0.05, one-sided Fisher test). Focusing on activation outliers and annotated cancer driver genes, we found 43 associations between 37 CGC cancer driver genes and 12 disease entities (Figure 5A). Some associations were already described in the literature (Supplementary Table 4). For example, we confirmed that the transcription factors *TLX1* and *TLX3* were associated with T-cell acute lymphoblastic leukemia, in line with previous reports (81,82). In addition, other studies mentioned the role of *HOXA11*, *PREX2,* and *RET* in AML (83–85). However, several associations have not yet been reported, including overexpression of *WNK2* in high-grade B-cell lymphoma, of *FAT4* in multiple myeloma, and of *LRP1B* in hairy cell leukemia (HCL), hairy cell leukemia variant (HCL-V) and marginal zone lymphoma (MZL).

**Figure 5.**
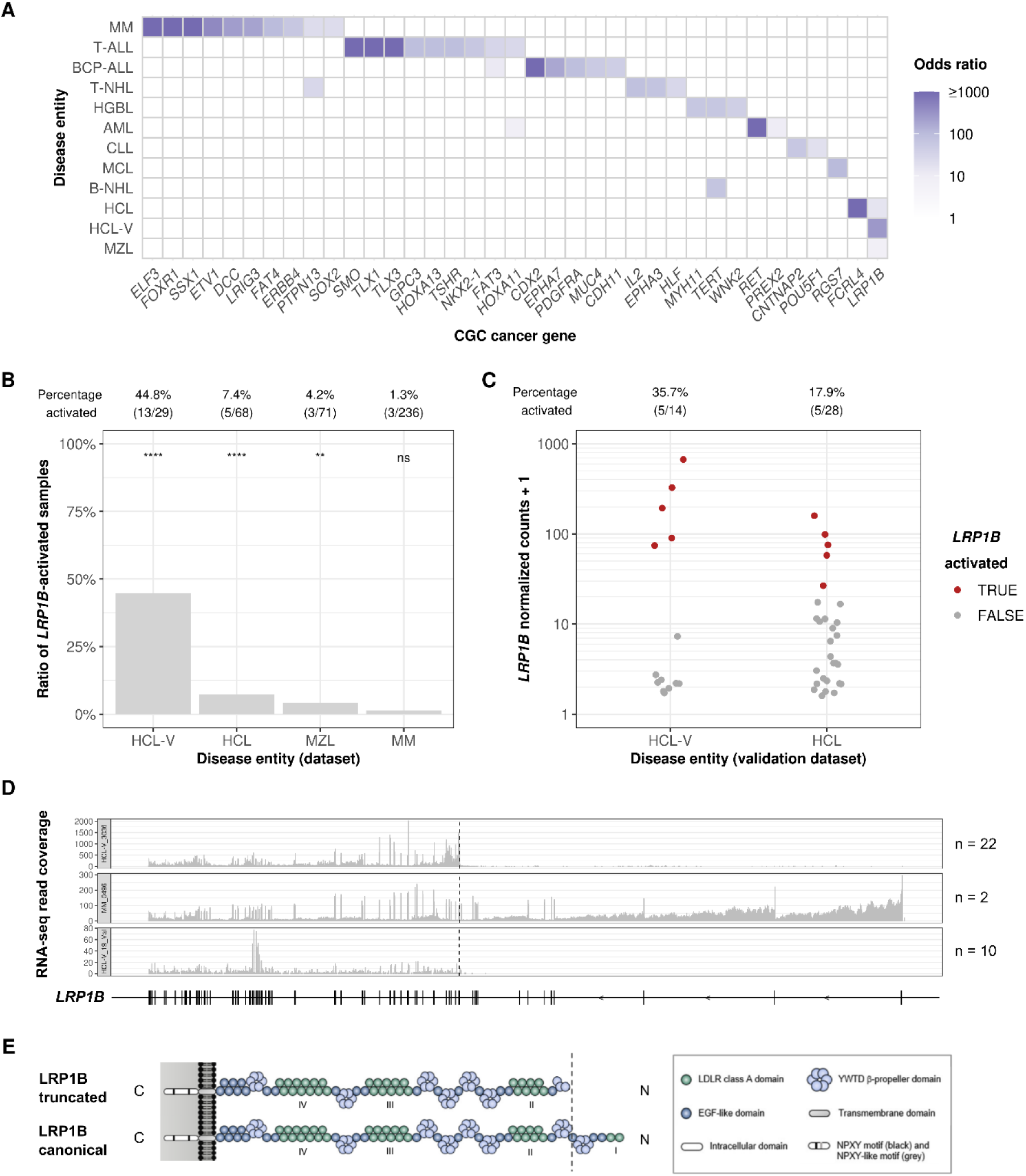
Aberrant activation of *LRP1B* is predominant in HCL-V. **(A)** Disease entities (full names in Table 1) against aberrantly activated CGC genes, colored by odds ratio from Fisher test. **(B)** Percentage of *LRP1B*-activated samples in the four disease entities in which *LRP1B* activation occurred. Percentages of samples showing *LPR1B* activation (NB-act) and nominal significance from the one-sided Fisher test are labeled at the top. **(C)** Normalized counts of *LRP1B*-activated samples of the dataset with regard to the different disease entities in the validation dataset. Percentages of samples showing *LPR1B* activation (Gaussian mixture clustering) are labeled at the top. **(D)** Transcriptomic coverage tracks showing *LRP1B* truncated isoforms of samples HCL-V_3036 and MM_0496 (dataset) and HCL-V_19_Val (validation dataset). **(E)** Anticipated domain organization of truncated and canonical *LRP1B* (adapted from Príncipe et al.(86)).

Remarkably, *LRP1B* was associated with very rare entities. *LRP1B*, or Low-Density Lipoprotein Receptor-related protein 1B, is a frequently altered gene in multiple cancer types, but its exact role remains unclear (86). In total, we found 24 samples (0.6% of all 3,760 samples) with aberrant high expression of *LRP1B*. Thereof 21 were found within HCL-V, HCL, and MZL, where *LRP1B*-activated samples made up 44.8% (HCL-V), 7.4% (HCL), and 4.2% (MZL) of each entity (Figure 5B). The other three cases with high *LRP1B* expression were found within multiple myeloma (MM). Among all *LRP1B*-activated samples, more than half of the cases (13 out of 24) were found in HCL-V patients, indicating that aberrant *LRP1B* expression might play an important role in HCL-V. *LRP1B* activation did not show a significant difference in overall survival (OS) in HCL-V patients. However, a trend towards shorter OS of patients with *LRP1B* activation was observed (median OS: 3.4 vs. 6.3 years; *P =* 0.45, two-sided log-rank test; Supplementary Figure 18). We then evaluated *LRP1B* expression in an independent validation dataset of 42 samples. Consistent with observations made on our primary dataset, we detected 10 *LRP1B*-activated samples in HCL-V (5/14; 36%) and HCL (5/28; 18%, Figure 5C) in the validation dataset. Moreover, the RNA-seq coverage for both datasets showed that samples overexpressing *LRP1B* expressed a truncated isoform in the majority (32/34; 94%) of samples (22/24 in the dataset; 10/10 in the validation dataset; Figure 5D, Supplementary Figure 19-20). The two samples expressing full-length transcripts were both multiple myeloma, whose causes are yet to be understood. In the truncated isoform cases, *LRP1B* expression started at exon 13, thus lacking the first 636 amino acids of exon 1 to 12. Three start codons located within exon 13 could enable the start of translation using the canonical open reading frame. Assuming this transcript is translated, it yields a truncated LRP1B protein starting from the middle of the second β-propeller domain (Figure 5E). However, we could not pinpoint a genomic cause for the truncated isoform, as no single nucleotide variants, short insertions or deletions, structural variants, or gene fusions involving *LRP1B* were found to be specific to the affected samples.

## Discussion

The study of rare cancer aberrations is an emerging field that greatly benefits from the growing large-scale next-generation sequencing (87,88). We have generated an extensive census of rare genomic and transcriptomic aberrations across 3,760 patients spanning 24 hematologic malignancy disease entities. This census is based on the largest collection of hematologic malignancy samples that have undergone WGS along with matched RNA-seq data, which also includes rare disease entities. We cannot share without access restrictions the exact variants, expression outliers, and splicing outliers at the sample level, as such data would compromise research participant privacy. Nevertheless, we publicly provide our census (Availability of data and materials) and have demonstrated its utility. This census comprises frequencies of genes harboring variants predicted to cause aberrant splicing, expression outliers, and splicing outliers. All these categories were significantly enriched for known hematologic malignancy driver genes, highlighting their role as putative drivers in the corresponding samples. Notably, we have reaffirmed the well-established associations between transcription factors *TLX1* and *TLX3* with T-cell acute lymphoblastic leukemia, as well as *HOXA11*, *PREX2*, and *RET* with AML. Furthermore, we built pan-leukemia and entity-specific driver gene predictors by integrating this data which successfully recovered known drivers and yielded promising novel candidates.

One of our notable findings is the identification of *LRP1B* as a potential marker for a subgroup of HCL-V and related B-cell malignancies. In our RNA-seq samples, *LRP1B* expression was rarely detected, occurring in only approximately 1% of cases. However, it was highly expressed in some cases of MZL, HCL, and approximately 50% of HCL-V, which we confirmed in an independent dataset. These three disease entities are mature B-cell malignancies and were previously regarded as separate entities in the revised 4^th^ edition of the WHO classification of hematolymphoid tumors, distinguished based on immunophenotypic markers and molecular genetics (89). HCL-V is typically resistant to conventional HCL therapy and does not show the HCL-specific *BRAF*-V600E mutation. However, as HCL, HCL-V, and MZL arise from malignant mature B-cells showing similar morphology, clear discrimination using conventional diagnostic techniques is often not possible. Thus, in the recently published 5^th^ edition of WHO classification, the term “HCL-V’’ has been removed, recognizing that the biology of this disease is unrelated to HCL (3). Instead, these cases are now considered splenic B-cell lymphoma/leukemia with prominent nucleoli (SBLPN), which also comprises rare cases of splenic MZL and B-prolymphocytic leukemia based on similar cytomorphological features. SBLPN rather serves as a placeholder for those morphologically defined cases of B-cell lymphoma not being classifiable into biologically distinct entities based on current evidence-based knowledge. We observed a tendency for a worse prognosis for HCL-V samples expressing *LRP1B*, though a larger sample size is needed to establish a statistical significance. Overall, our results suggest a potential subcategorization of HCL-V/SBLPN based on *LRP1B* expression, whose functional implications remain to be elucidated.

*LRP1B*, also known as Low-Density Lipoprotein Receptor-related protein 1B, is broadly expressed in multiple normal tissues but not in blood or bone marrow (90–92). It plays a role in various biological processes such as angiogenesis, chemotaxis, proliferation, adhesion, apoptosis, endocytosis, immunity, host-virus interaction, and protein folding. Additionally, *LRP1B* is among the most frequently altered genes in human cancer overall (93–98). For tissues where *LRP1B* is normally expressed, *LRP1B* is often inactivated in cancer through several genetic and epigenetic mechanisms, making it a putative tumor suppressor gene. Overexpression of a truncated isoform of *LRP1B* in cells that normally do not express it, as we observed here, could play a role in oncogenesis by disrupting similar biological processes. Despite the challenges posed by the enormous size of LRP1B, as highlighted in the study by Príncipe et al. (86), our findings encourage further investigations to unravel the potential role of *LRP1B* in B-cell malignancies.

Our study has limitations. Our approach is not geared towards retrieving common abnormalities and may underscore non-regulatory mutations. For example, *NPM1*, one of the most frequently mutated genes in AML (99), was not prioritized. *NPM1* was categorized as a driver gene by two intOGen tools but not by the other five. Pathogenic mutations of *NPM1* act post-translationally by affecting the cellular localization of the NPM1 protein. Not surprisingly, the transcriptome data of *NPM1* were not informative. Consequently, *NPM1* was not prioritized by our driver gene prediction model. Our dataset, which is derived from routine diagnostic sampling, does not include matched healthy controls. Therefore, the variants we considered probably include more rare somatic variants than in studies with matched control samples. Additionally, we did not include gene fusion and copy number variation calls, which are frequent causes of hematologic malignancies, into the driver gene prediction model, as we found them to have very high false positive rates during preliminary investigations. Moreover, we have purposely decided to restrict the input data of the driver gene prediction model to the sole genomic and transcriptomic data of our dataset in order to provide the community predictions unbiased by previous literature. Future work could integrate this resource as prior information on pathways or protein interaction networks or with complementary datasets to provide a refined landscape of genomic and transcriptomic aberrations driving hematologic malignancies.

## Conclusions

We established a unique and comprehensive census encompassing the genomic and transcriptomic landscape of 3,760 hematologic malignancy samples, covering a wide range of disease entities. This comprehensive census can be leveraged to identify novel biomarkers, propose therapeutic decisions, and unravel the molecular underpinning of the heterogeneity of hematologic cancers.

## Declarations

## Ethics approval and consent to participate

All patients or their legal guardians gave written informed consent for genetic analyses and to the use of laboratory results and clinical data for research purposes, according to the Declaration of Helsinki. The study was approved by the MLL’s institutional review board.

## Consent for publication

Not applicable.

## Availability of data and materials

The census of genomic and transcriptomic aberrations of 3,760 hematologic malignancy samples spanning 24 disease entities is available at https://zenodo.org/record/8341457. A data transfer agreement must be obtained to access further data, including raw genomic and transcriptomic sequencing data, as they could compromise research participant privacy. To this end, contact St.H. at MLL.

All code is available at https://github.com/gagneurlab/Leukemia_outlier.

## Authors’ contributions

L.W., St.H., and J.G. designed the research; X.C., Sa.H., A.J.A., M.S., P.S., and St.H. performed the research; C.O., and St.H. collected the data; X.C., Sa.H., A.J.A., M.S., I.S., and N.W. analyzed and interpreted the data; X.C., Sa.H., and S.V. performed statistical analysis; X.C., and A.J.A. developed the software; M.H., L.W., St.H., and J.G. supervised the research; M.H., T.H., St.H., and J.G. acquired the funding; X.C., Sa.H., A.J.A., F.R.T., M.S., S.V., L.W., St.H., and J.G. wrote the manuscript; and all authors reviewed and agreed to the final version of the manuscript.

## Supporting information

Supplementary Material and Methods, Table description

## Data Availability

All data produced in the present study are available upon reasonable request to the corresponding authors.

https://zenodo.org/record/8341457

## Acknowledgments

The authors thank everyone from the MLL 5k project involved in patient inclusion, sample collection, and sequencing data generation; everyone involved in patient inclusion, sample collection, and sequencing data generation of the HCL-V validation dataset; Stefan Loipfinger, Luise Schuller, and Kim Anh Lilian Le for their contributions to the initial work of this project; Felix Brechtmann and Christian Mertes for the enlightening discussions.

## Authors’ information

Correspondence: Julien Gagneur, School of Computation, Information and Technology, Technical University of Munich, Boltzmannstraße 3, 85748 Garching bei München, Germany; and Stephan Hutter, Münchner Leukämielabor GmbH (MLL), Max-Lebsche-Platz 31, 81377 München, Germany; and Leonhard Wachutka, School of Computation, Information and Technology, Technical University of Munich, Boltzmannstraße 3, 85748 Garching bei München, Germany.

## List of abbreviations

CGC: Cancer Gene Census
LRP1B: Low-Density Lipoprotein Receptor-related protein 1B
MLL: Munich Leukemia Laboratory
NB-act: Negative Binomial activation
RNA-Seq: total RNA sequencing
VEP: Ensembl variant effect predictor
WGS: Whole Genome Sequencing

