## Supplementary Material and Methods, Table description for "Analysis of 3,760 hematologic malignancies reveals rare transcriptomic aberrations of driver genes"

### Supplementary Materials and Methods

DNA sequencing, read mapping, variant calling, and copy number variations calling

Whole genome sequencing (WGS) libraries were prepared from 1µg of DNA with the TruSeq PCR free library prep kit following the manufacturer's recommendations (Illumina, San Diego, CA, USA) and 2x150bp paired-end sequences were generated on a NovaSeq 6000 or HiSeqX instrument with 100x coverage (Illumina, San Diego, CA, USA). Reads were aligned to the human reference genome (GRCh37, Ensembl annotation) using the Isaac aligner (v3.16.02.19) (1) through BaseSpace's WGS app (v5, Illumina, San Diego, CA, USA) with default parameters. Resulting BAM files were used in BaseSpace's Tumor/Normal app (v3) to call single nucleotide variants and short insertions and deletions (<50bp) with Strelka (v2.4.7) (2) and large-scale structural variants with Manta (v0.28.0) (3). As no sample-specific normal tissue was available, a so-called unmatched normal was used in its place to reduce technical artifacts and germline calls. For this, WGS was performed on gender-matched genomic DNA from a mixture of multiple anonymous donors (Promega, Fitchburg, WI, USA). Copy number variations were called with GATK (v4.0.8.1) (4) using the Broad Institute's recommended best practices pipeline. Here, two panels of normals (PON) were used for denoising, consisting of 124 female and 191 male samples, which presented a normal karyotype during routine diagnostics.

RNA sequencing, read mapping, and fusion detection

TruSeq Total Stranded RNA kit was used, starting with 250 ng of total RNA, to generate RNA libraries following the manufacturer's recommendations (Illumina, San Diego, CA, USA). 2×100 bp paired-end reads were sequenced on the NovaSeq 6000 (Illumina, San Diego, CA, USA) with a median of 50 million reads per sample. Using BaseSpace's RNA-seq Alignment app (v2.0.1) with default parameters, reads were mapped with STAR aligner (v2.5.0a) (5) to the human reference genome hg19 (RefSeq annotation). Single nucleotide variants and short insertions and deletions were called using Isaac variant caller (v2.3.13)(1). Fusion calling was performed with Manta (v0.29.0)(3), Arriba (v1.2.0) (6), and STAR-Fusion (v1.9.0) (7). Only fusions that were called by at least two algorithms were considered in order to reduce false positives.

<u>NB-act</u>

NB-act (Negative Binomial activation) estimates the probability of a gene that is generally not expressed in the population to be expressed in a particular sample. It was applied to the rarely-expressed genes of all samples from the dataset.

We assumed that the observed count $k_{ij}$ of gene $i = 1,..., p$ in sample $j = 1,..., m$ followed a negative binomial distribution with a shared dispersion parameter $\theta$ and expected value $\mu_{ij}$:

$$P(k_{ij}) = NB(k_{ij}|\mu_{ij}, \theta)$$

The dispersion parameter $\theta$ was set to 0.02 and corresponds to the empirically observed lowest dispersion values estimated by OUTRIDER on expressed genes. As dispersion inversely relates to variance, this value makes NB-act conservative. OUTRIDER uses 1 fragment per kilobase of transcript per million mapped reads (FPKM) as the upper expression limit for a lowly-expressed gene. Therefore, we set the expected count $\mu_{ij}$ of a gene is given as the product of 1 FPKM, the sample-specific size factor $s_j$, and the exon length of the gene $L_i$ [nt] divided by 1,000 to get absolute count expectations:

$$\mu_{ij} := 1 * s_j * L_i / 1000$$

The size factors $s_j$ capture variations in sequencing depth and were estimated using DESeq2 (8). The exon length $L_i$ [nt] was extracted from Gencode v33b. The *P*-values were calculated and corrected for multiple testing using functions provided by OUTRIDER (‘alternative=greater’). All significant outliers of this analysis were named “activation outliers”.

CGC

The Cancer Gene Census (CGC) is a catalog of those genes that contain mutations that have been causally implicated in cancer and explains how the dysfunction of these genes drives cancer (9). The CGC genes GRCh37 v97 were obtained from https://cancer.sanger.ac.uk/census and subsetted for the subsequent analysis, including enrichment, association, and machine learning model (Table 2).

Table 2. CGC subsetting criteria

| Subset | Column ‘Tissue Type’ | Column ‘Role in Cancer’ | Number of genes |
|---|---|---|---|
| CGC cancer driver gene | any | any | 721 |
| CGC cancer oncogene | any | containing ‘oncogene’ | 314 |
| CGC cancer tumor suppressor gene | any | containing ‘TSG’ | 318 |
| CGC hematologic malignancy driver gene | containing ‘L’* | any | 322 |
| CGC hematologic oncogene | containing ‘L’* | containing ‘oncogene’ | 156 |
| CGC hematologic tumor suppressor gene | containing ‘L’* | containing ‘TSG’ | 134 |

*L: leukemia or lymphoma

The cancer type of the gene was determined by the ‘Tissue Type’ annotation. All genes were labeled as ‘cancer’. Genes whose ‘Tissue Type’ included ‘L’ (leukemia or lymphoma) were further labeled as ‘leukemia’. The role of the gene was determined by the ‘Role in Cancer’ annotation. Genes whose ‘Role in Cancer’ included ‘oncogene’ or ‘TSG’ were labeled as ‘oncogene’ or ‘tumor suppressor gene’ respectively. Some genes have been reported with both roles and can have received both labels.

Hematologic panel genes

The gene panel consists of genes that are required for diagnosis and have been found to be associated with prognosis or are relevant for therapy in hematologic malignancies (10,11). The gene panel furthermore contains genes that have been described in the literature as being recurrently mutated in one or more hematologic disease entities. Altogether, these were 122 genes, partially overlapping CGC hematologic malignancy driver genes. These 122 genes were used for curation, sample genotyping analysis, and machine learning models:

*APC, ARID1A, ASXL1, ASXL2, ATM, ATRX, BCL2, BCOR, BCORL1, BIRC3, BRAF, BRCC3, BTK, CALR, CARD11, CBL, CCND1, CD79A, CD79B, CDH23, CDKN2A, CEBPA, CHEK2, CREBBP, CSF3R, CSNK1A1, CTCF, CUX1, CXCR4, DDX3X, DDX41, DDX54, DHX29, DIS3, DNMT3A, EP300, ETNK1, ETV6, EZH2, FAM46C, FANCL, FAS, FAT4, FBXW10, FBXW7, FLT3, FOXO1, GATA1, GATA2, GNAS, GNB1, GPR98, ID3, IDH1, IDH2, IKBKB, IL2RG, JAK1, JAK2, JAK3, KDM5A, KDM6A, KIT, KLF2, KLHL6, KMT2D, KRAS, LRP1B, MAP2K1, MAPK1, MEF2B, MPL, MYBBP1A, MYC, MYD88, NF1, NFKBIE, NOTCH1, NOTCH2, NPM1, NRAS, PHF6, PIGA, PLCG2, POT1, PPM1D, PRPF8, PTPN11, PTPRD, RAD21, RB1, RPS15, RUNX1, SETBP1, SF1, SF3A1, SF3B1, SH2B3, SMC1A, SMC3, SRSF2, STAG2, STAT3,*

*STAT5B, SUZ12, TBL1XR1, TCF3, TET2, TLR2, TNFAIP3, TNFRSF14, TP53, TRAF3, U2AF1, U2AF2, UBR5, WHSC1, WT1, XPO1, ZBTB7A, ZMYM3, ZRSR2*

#### Driver gene prediction

##### Features

For each gene, the result of each tool was summarized as numerical values, which reflected its mutation, expression, and splicing aberrations profile. Therefore, all genes were assigned with vectors of numbers, resulting in a feature matrix (genes x features). The feature vectors were generated for each of the tool in two ways:

1) Per study group: using only samples from each of the 14 study groups
2) Complete dataset: using all samples

##### OUTRIDER features

For each study group and for the complete dataset, the 22 OUTRIDER features per gene were generated, which consisted of:

1) The number of samples for which the gene is among the top 1, 5, 10, 25, 50 overexpression outliers sorted by False Discovery Rate (FDR) on the one hand and underexpression outlier on the other hand (10 features)
2) The number of samples for which the gene is among the significant overexpression outliers for FDR cutoffs of 0.01, 0.05, 0.1 on the one hand and among underexpression outliers on the other hand (6 features)
3) The number of samples for which the gene z-score is larger than 2, 4, or 6 on one hand and less than -2, -4, -6 on the other hand (6 features)

#### NB-act features

For each study group and for the complete dataset, the 11 NB-act features per gene were generated, which consisted of:

1) The number of samples for which the gene is among the top 1, 5, 10, 25, 50 overexpression outliers sorted by FDR (5 features)
2) The number of samples for which the gene is among the significant overexpression outliers for FDR cutoffs of 0.01, 0.05, 0.1 (3 features)
3) The number of samples for which the gene z-score is larger than 2, 4, or 6 (3 features)

#### FRASER features

For each study group and for the complete dataset, the 22 FRASER features per gene were generated, which consisted of:

1) The number of samples for which the gene is among the top 1, 5, 10, 25, 50 overrepresented splicing outliers sorted by FDR on the one hand, and underrepresented splicing outlier on the other hand (10 features)
2) The number of samples for which the gene is among the significant overrepresented splicing outliers for FDR cutoffs of 0.01, 0.05, 0.1 on the one hand and among underrepresented splicing outliers on the other hand (6 features)
3) The number of samples for which the gene delta Intron Jaccard Index is larger than 0.1, 0.2, 0.3 on the one hand, and less than -0.1, -0.2, -0.3 on the other hand (6 features)

#### AbSplice features

For each study group and for the complete dataset, the 9 AbSplice features were generated per gene, which consisted of:

1) The maximum AbSplice-DNA score across all rare variants and samples by cutoffs 0.01, 0.05, 0.2 (3 features)

2) The mean AbSplice-DNA score across all rare variants and samples by cutoffs 0.01, 0.05, 0.2 (3 features)
3) The number of samples for which the gene has at least one rare variant with an AbSplice-DNA score larger than 0.01, 0.05, 0.2 (3 features)

##### intOGen features

The intOGen feature vectors were generated separately from the result of the seven individual methods. The evaluation metrics of the seven methods were chosen the same way as intOGen and then transformed accordingly (Table 3). The mode of action (e.g., role as an oncogene or tumor suppressor gene) was calculated the same way as intOGen. Each transformed metric was assigned into the corresponding category ‘Activating’, ‘Loss-of-function’, or ‘ambiguous’ based on the mode of action, resulting in 7*3=21 features.

Table 3. Seven methods and the corresponding metric, filter, and transformation

| Method | Metric and transformation | Filter |
|---|---|---|
| HotMAPS (12) | -log(q-value) | - |
| OncodriveCLUSTL (13) | sum(SCORE) | - |
| smRegions (14) | max(U) | - |
| OncodriveFML (15) | -log(Q_VALUE) | - |
| MutPanning (16) | -log(FDR) | Significance < 0.1 |
| dNdScv (17) | -log(qallsubs_cv) | - |
| CBaSE (18) | -log(q_pos) | - |

#### Random forest classifier

The random forests were trained using the function ‘RandomForestClassifier’ with ‘n_estimators’ = 100, ‘criterion’ = ‘gini’, ‘max_depth’ = 10, ‘min_samples_split’ = 19, ‘min_samples_leaf’ = 1, ‘min_weight_fraction_leaf’ = 0, ‘max_features’ = ‘auto’, ‘max_leaf_nodes’ = None, ‘min_impurity_decrease’ = 0, ‘bootstrap’ = True, ‘oob_score’ = False’, ‘random_state’ = None, ‘verbose’ = 0, ‘warm_start’ = False‘, class_weight’ = None, ‘ccp_alpha’ = 0, and ‘max_samples’ = None.

#### Supplementary Data

Supplementary Table

Supplementary Table 1. Prediction result of the hematologic malignancy drive gene prediction model using the complete dataset

Supplementary Table 2. Prediction result of the hematologic malignancy driver gene prediction model using each of the 14 study groups

Supplementary Table 3. Associations identified between disease entities and genes

Supplementary Table 4. Curation of associations between CGC cancer driver genes disease entities using Activation outliers

Supplementary Figure

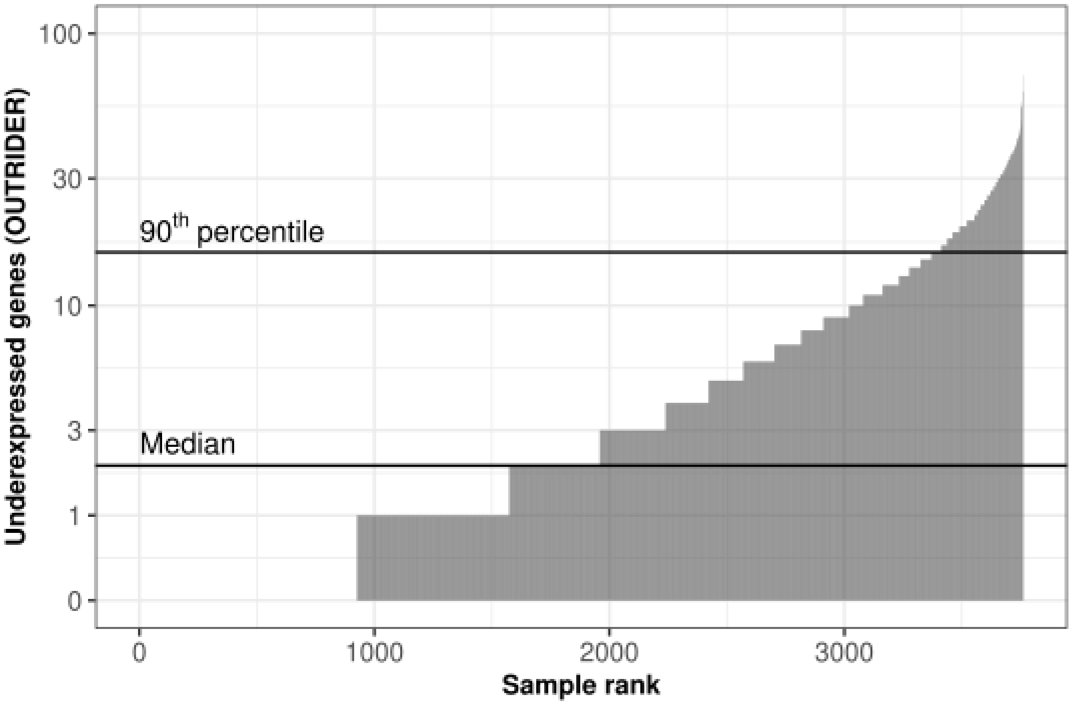


**Supplementary Figure 1. Underexpressed genes (OUTRIDER) per sample.** Horizontal lines mark the median and the 90th percentile.

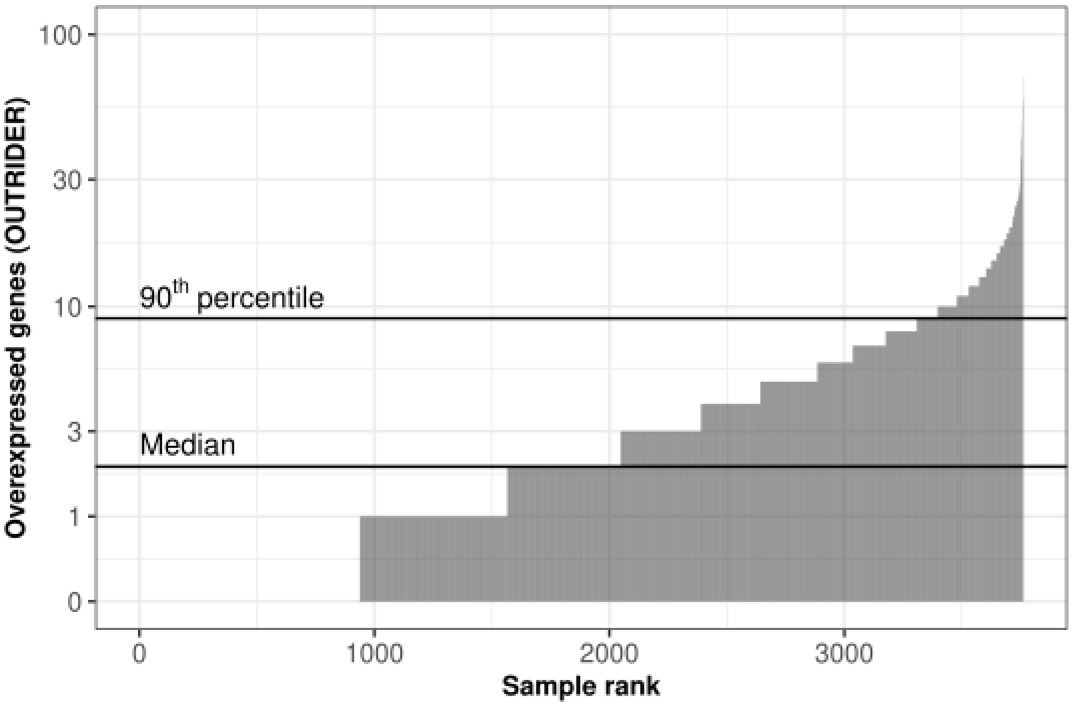


**Supplementary Figure 2. Overexpressed genes (OUTRIDER) per sample.** Horizontal lines mark the median and the 90th percentile.

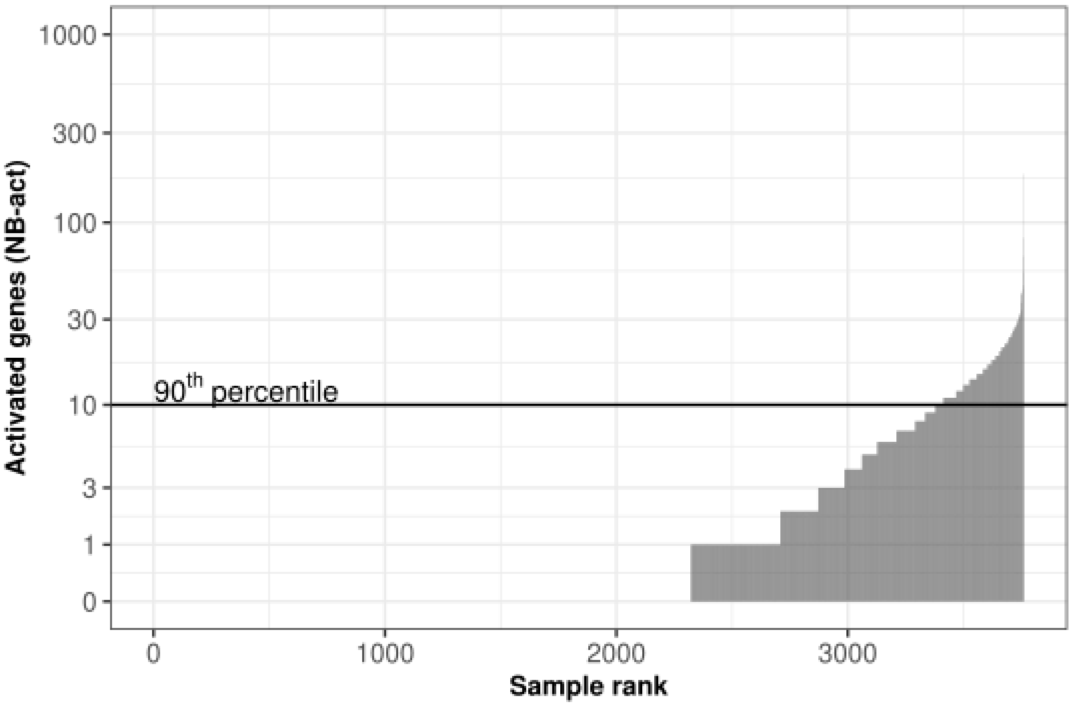


**Supplementary Figure 3. Activated genes (NB-act) per sample.** Horizontal line marks the 90th percentile.

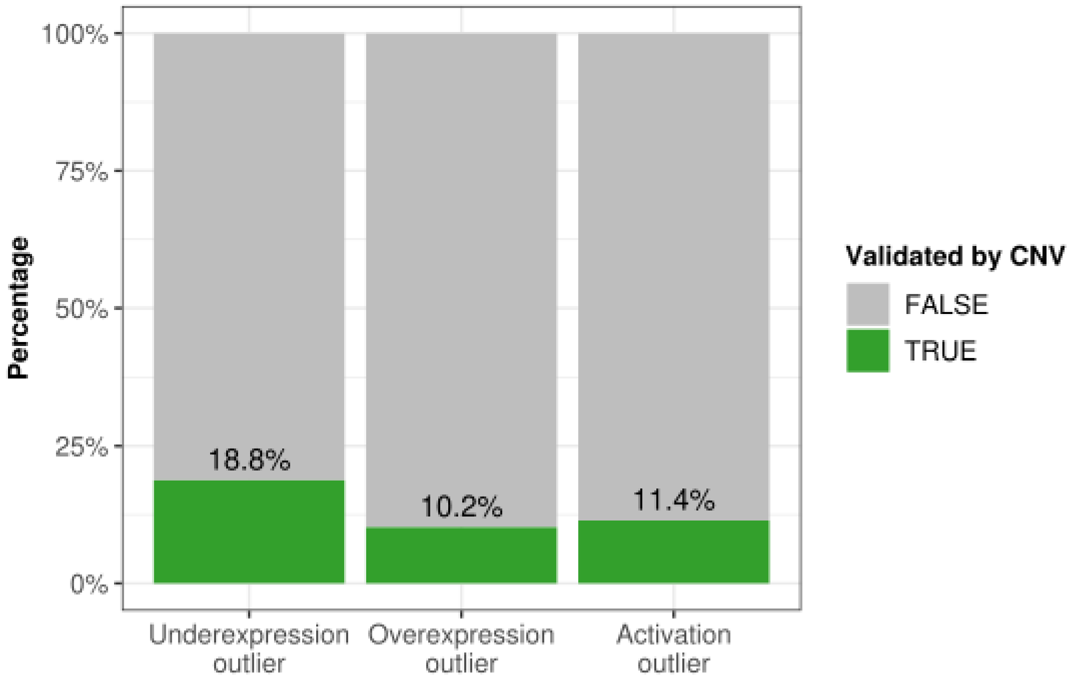


**Supplementary Figure 4.** Expression outliers (OUTRIDER, NB-act) are explained by copy number variations (GATK) among the different categories of expression outliers on the sample-gene level.

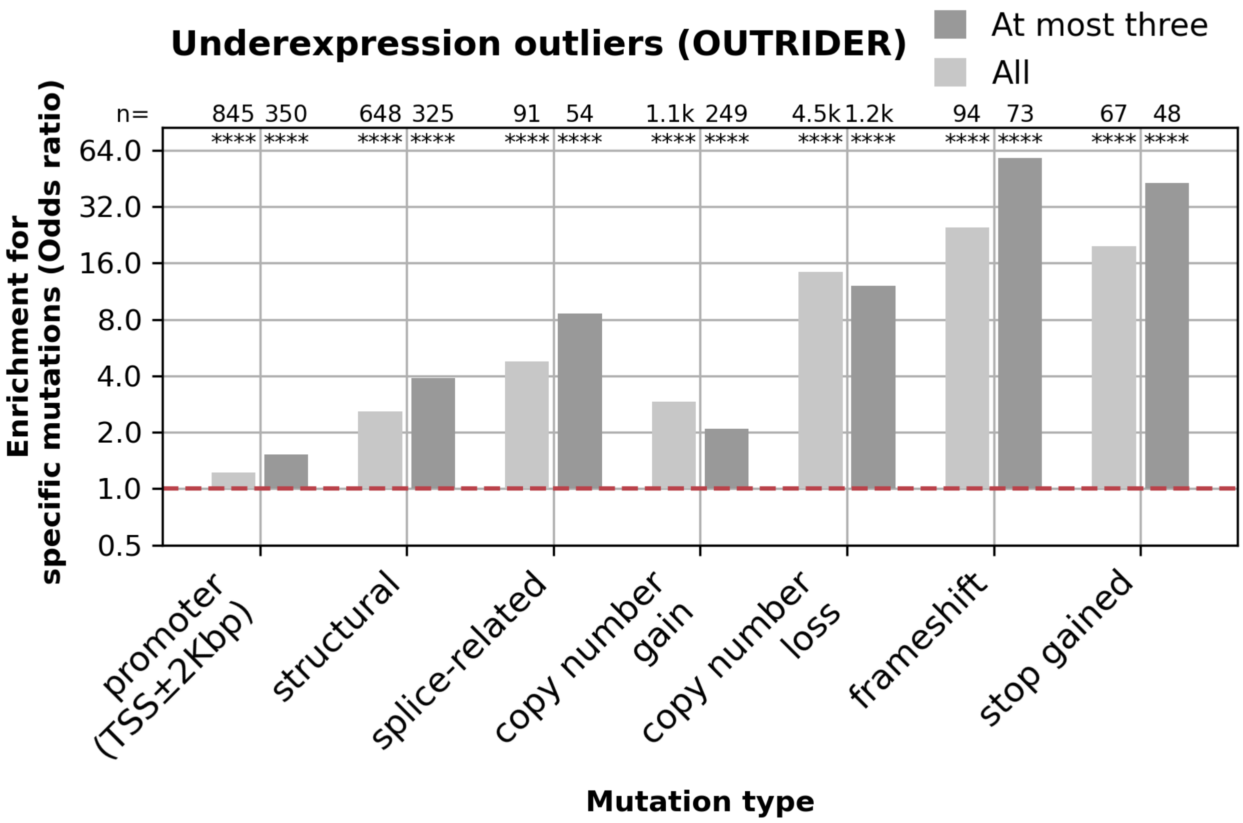


**Supplementary Figure 5.** Enrichment for different mutation types among all genes called by OUTRIDER as well as at most three significant genes per sample that were called to be underexpression outliers. Numbers of the mutations and nominal significances from the Fisher test are labeled at the top of the bars. (ns: not significant; *: P ≤ 0.05; **: P ≤ 0.01; ***: P ≤ 0.001; ****: P ≤ 0.0001)

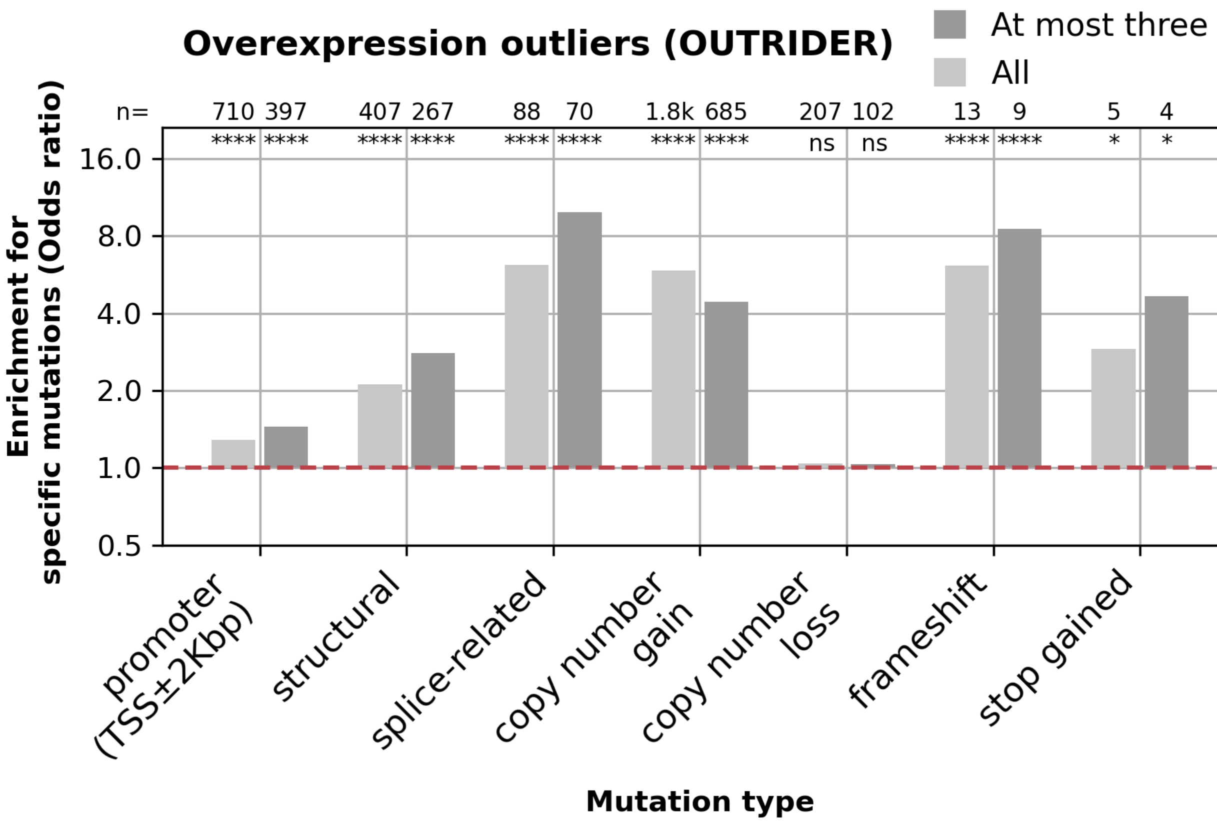


**Supplementary Figure 6.** Enrichment for different mutation types among all genes called by OUTRIDER as well as at most three significant genes per sample that were called to be overexpression outliers. Numbers of the mutations and nominal significances from the Fisher test are labeled at the top of the bars. (ns: not significant; *: P ≤ 0.05; **: P ≤ 0.01; ***: P ≤ 0.001; ****: P ≤ 0.0001)

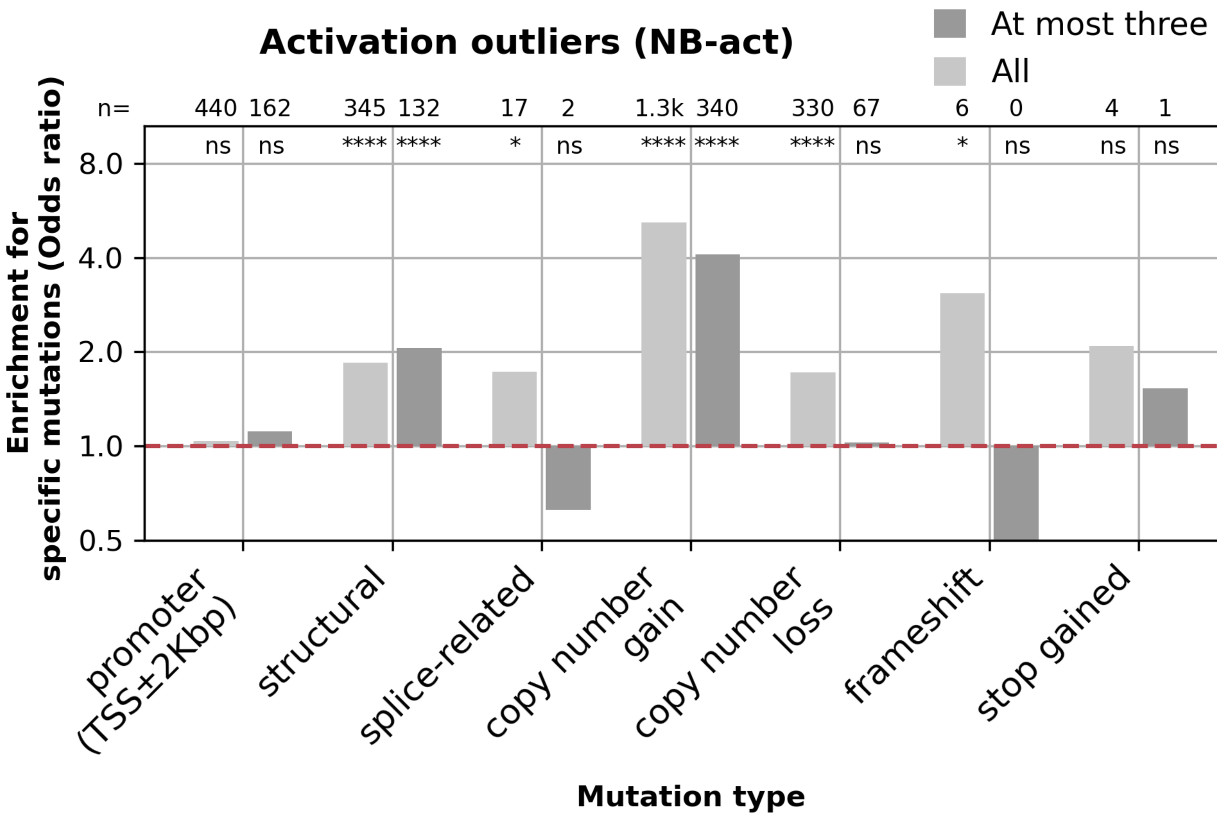


**Supplementary Figure 7.** Enrichment for different mutation types among all genes called by NB-act as well as at most three significant genes per sample that were called to be activation outliers. Numbers of the mutations and nominal significances from the Fisher test are labeled at the top of the bars. (ns: not significant; *: P ≤ 0.05; **: P ≤ 0.01; ***: P ≤ 0.001; ****: P ≤ 0.0001)

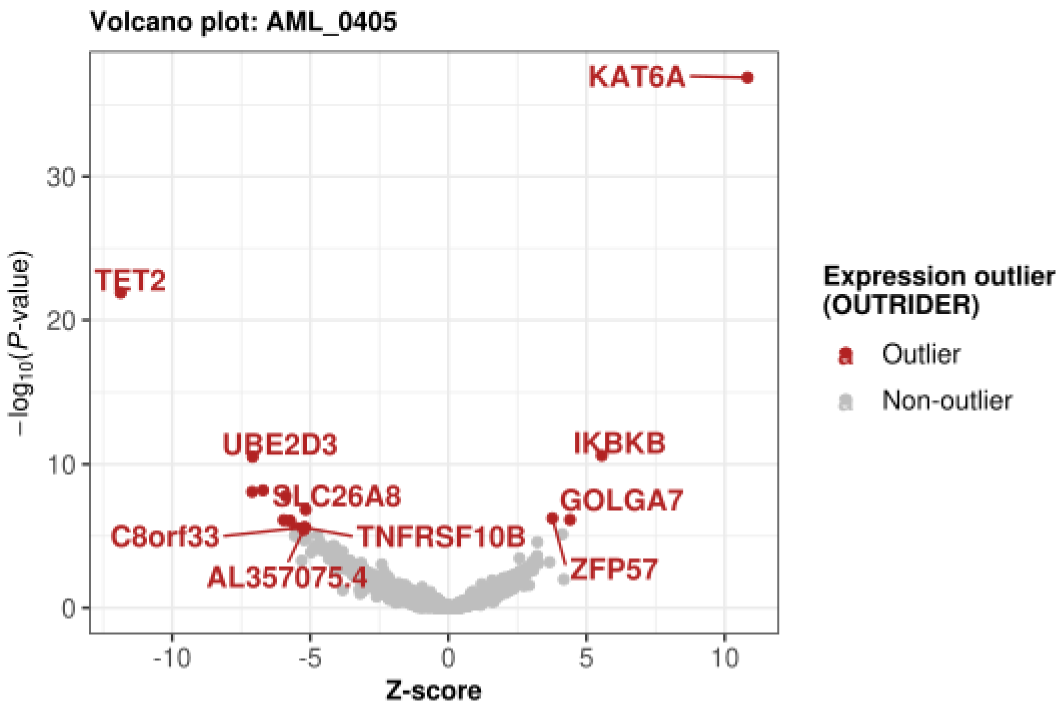


**Supplementary Figure 8. *P*-value against z-score of expression outliers in sample AML_0405.** Four overexpression outliers and 13 underexpression outliers (red) were detected.

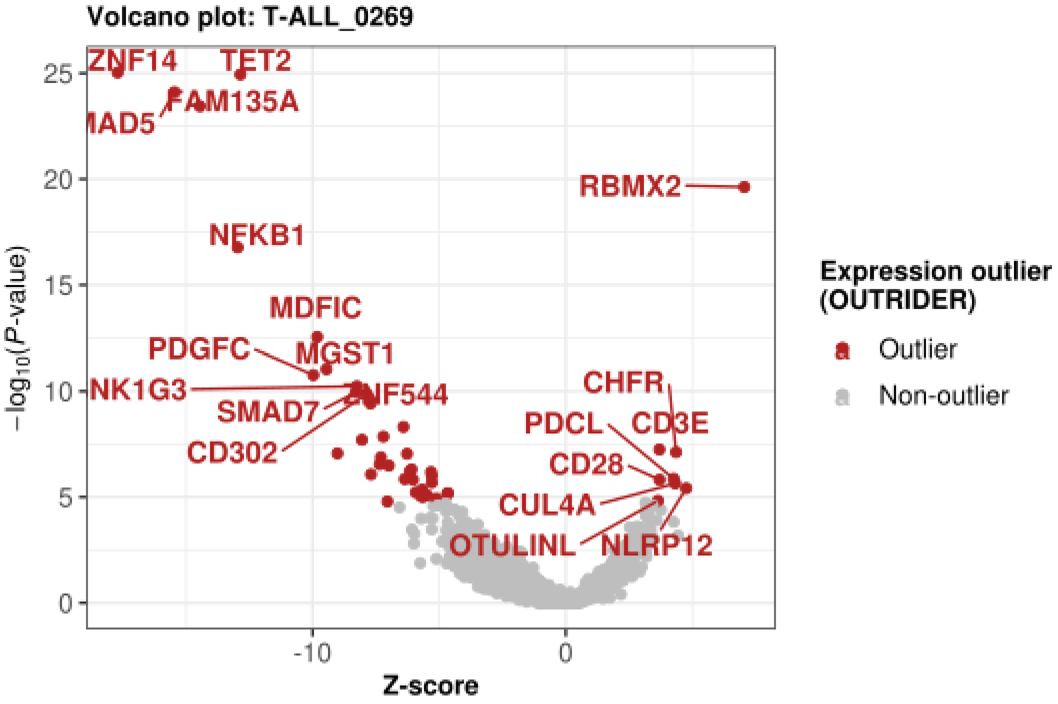


**Supplementary Figure 9. *P*-value and z-score of expression outliers in sample T-ALL_0269.** Eight overexpression outliers and 38 underexpression outliers were detected (red).

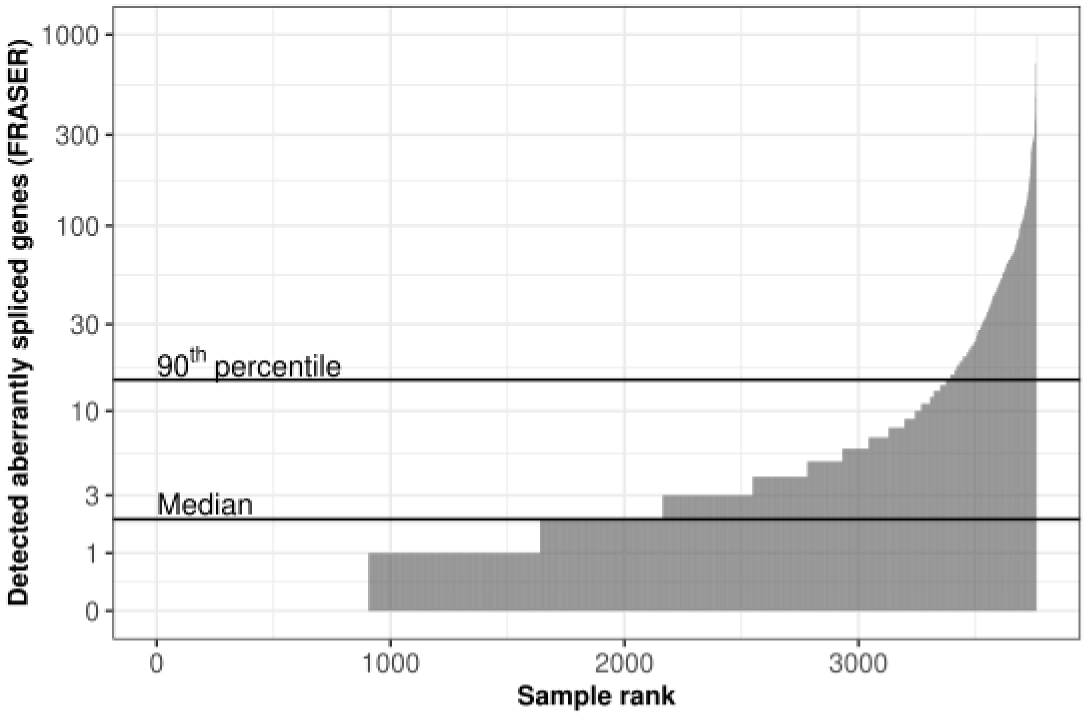


**Supplementary Figure 10. Aberrantly spliced genes (FRASER) per sample.** Horizontal lines mark the median and the 90th percentile.

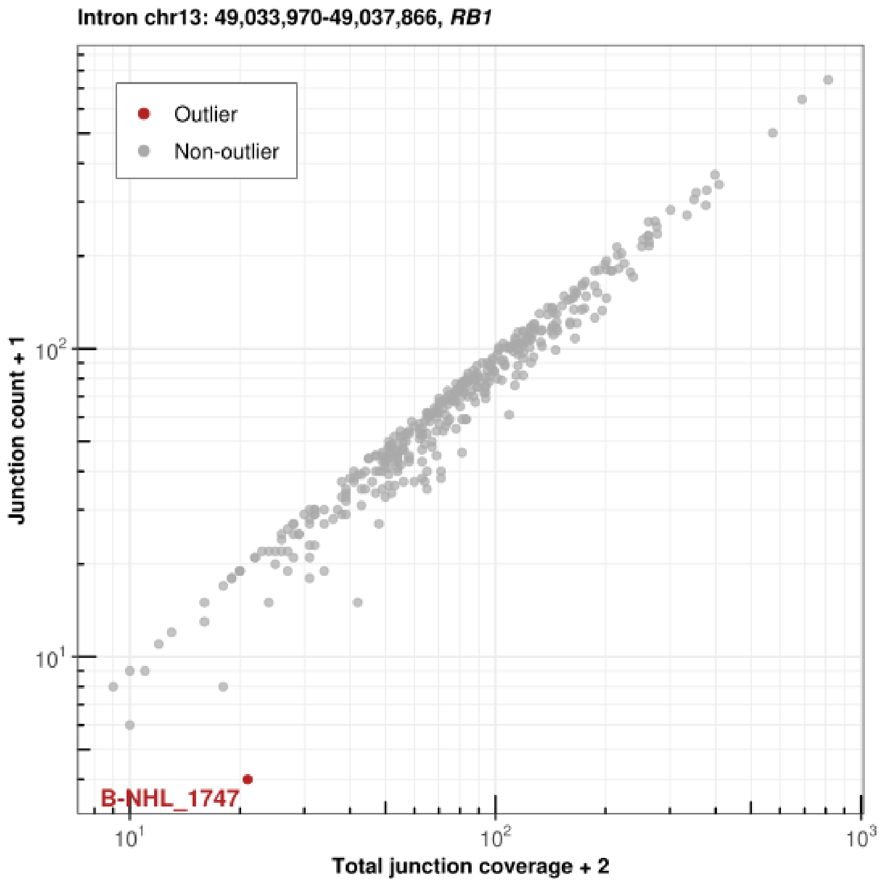


**Supplementary Figure 11. Junction counts (split reads) against the displayed intron's total junction coverage (exon-intron or intron-exon spanning reads) of *RB1* case study.** The displayed intron of the *RB1* only shows abnormal splicing in sample B-NHL_1747.

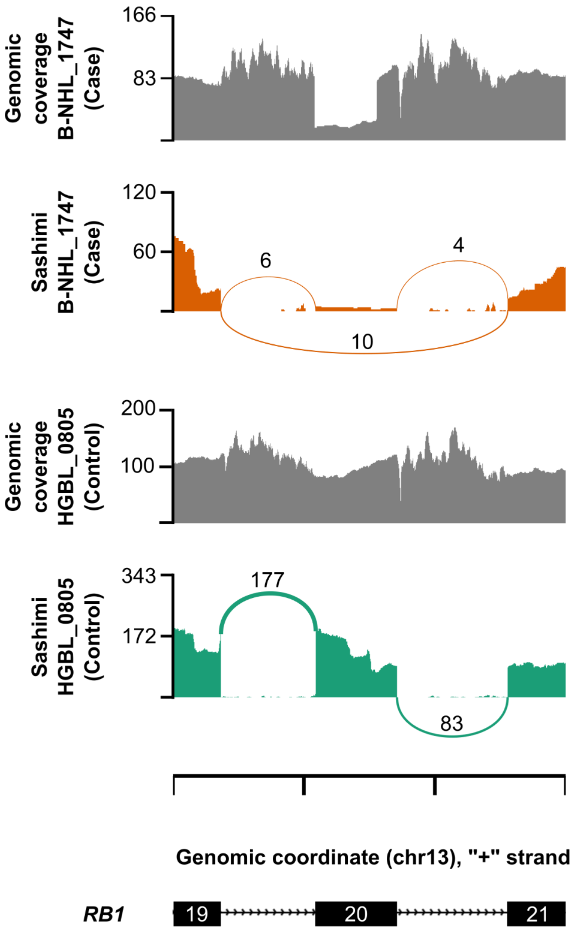


**Supplementary Figure 12. Genomic coverage and sashimi plots of *RB1* case study.** Genomic coverage of WGS (y-axis). Sashimi plots showing RNA-seq read coverage (y-axis) and the numbers of split reads spanning an intron indicated on the exon-connecting line for two aberrant splicing events. One case individual showing exon-skipping for *RB1* 20$^{th}$ and one control individual are displayed. The rare splice-affecting structural variant, which existed exclusively in the case individual, is observed in the genomic coverage plot as a drop of coverage.

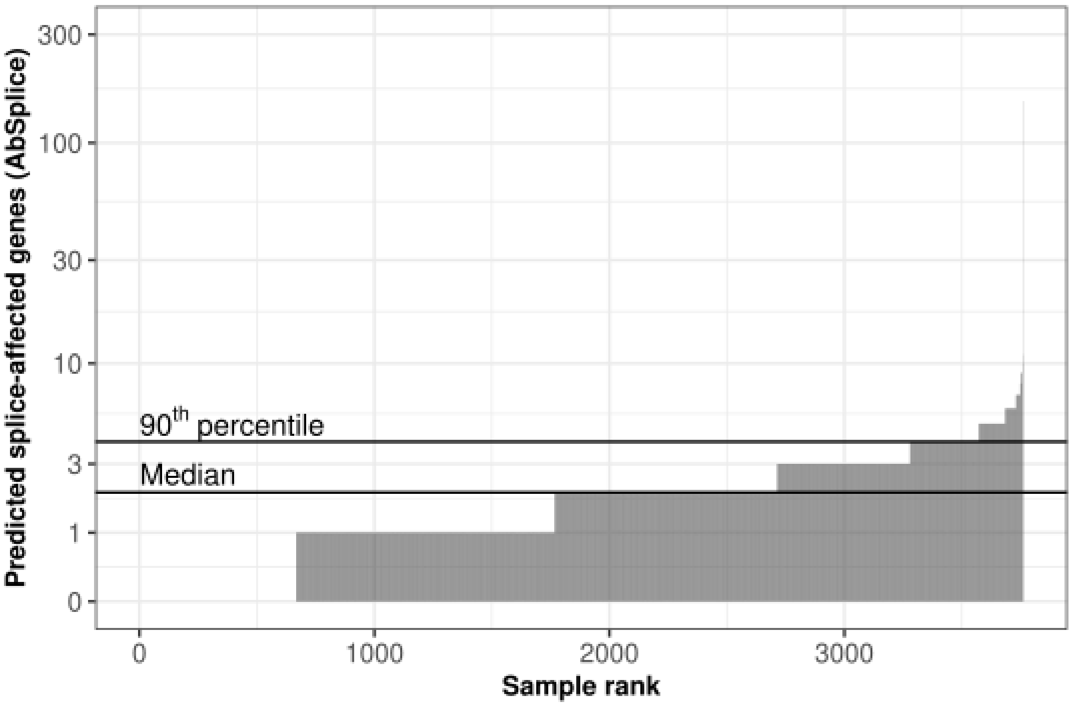


**Supplementary Figure 13. Predicted splice-affected genes (AbSplice) per gene.** Horizontal lines mark the median and the 90th percentile.

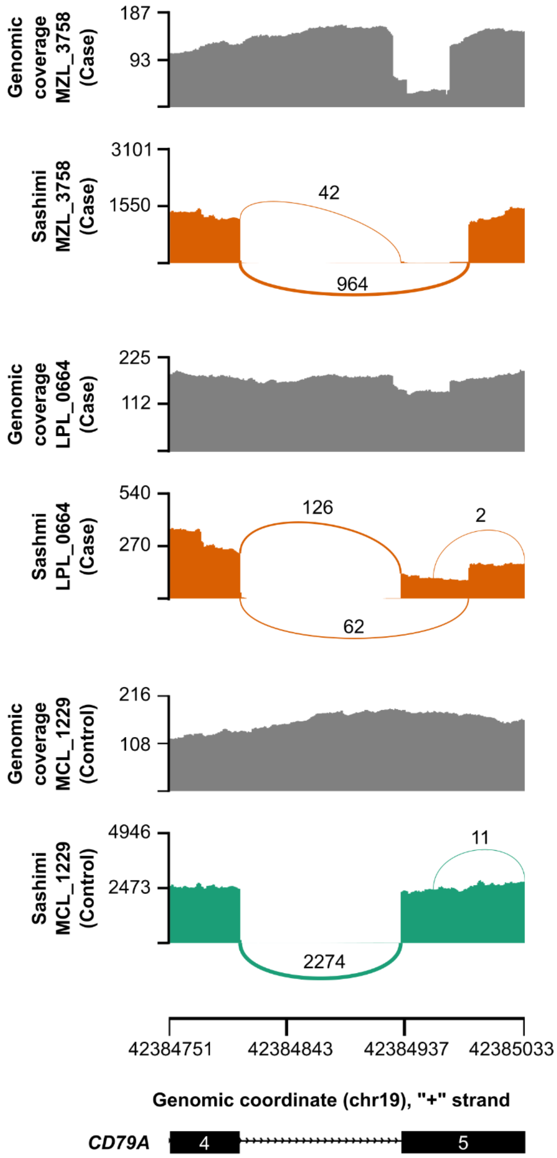


**Supplementary Figure 14. Genomic coverage and sashimi plots of *CD79A* case study.** Genomic coverage of WGS (y-axis). Sashimi plots showing RNA-seq read coverage (y-axis) and the numbers of split reads spanning an intron indicated on the exon-connecting line (using pysashimi) for two aberrant splicing events. Two case individuals using a unique splice-site in *CD79A* 5$^{th}$ exon acceptor site and one control individual are displayed. The rare splice-affecting deletion (NM_001783.4:c.568-2_610del) predicted by AbSplice, which existed exclusively in the two case individuals, is observed in the genomic coverage plot as a drop of coverage.

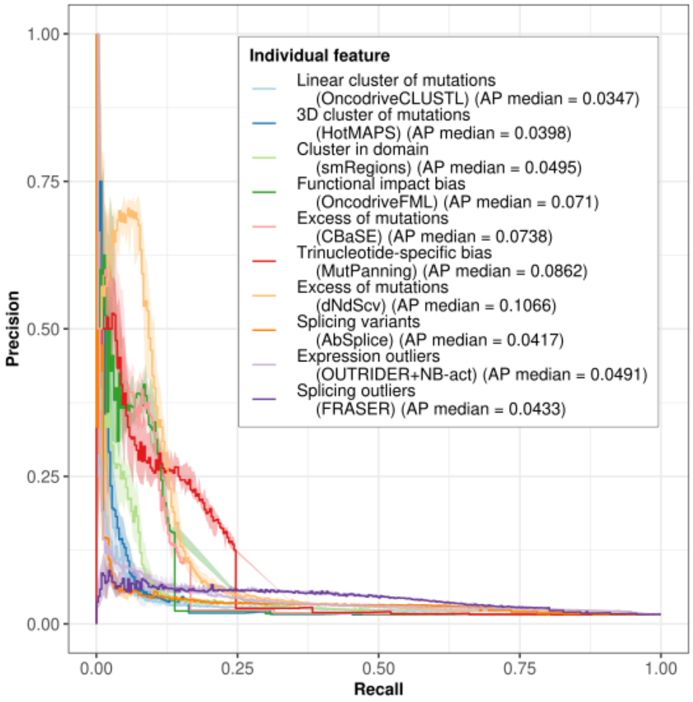


**Supplementary Figure 15. Precision recall curve showing performance of hematologic malignancy driver gene prediction model when using individual features and complete dataset.** Shading represents the confidence interval of 10 random repeats.

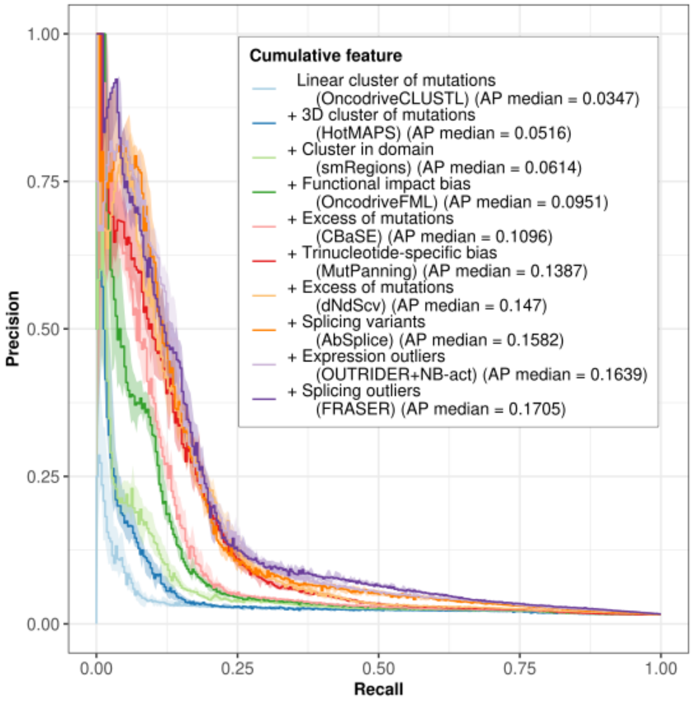


**Supplementary Figure 16. Precision recall curve showing performance of hematologic malignancy driver gene prediction model when using cumulative features and complete dataset.** Shading represents the confidence interval of 10 random repeats.

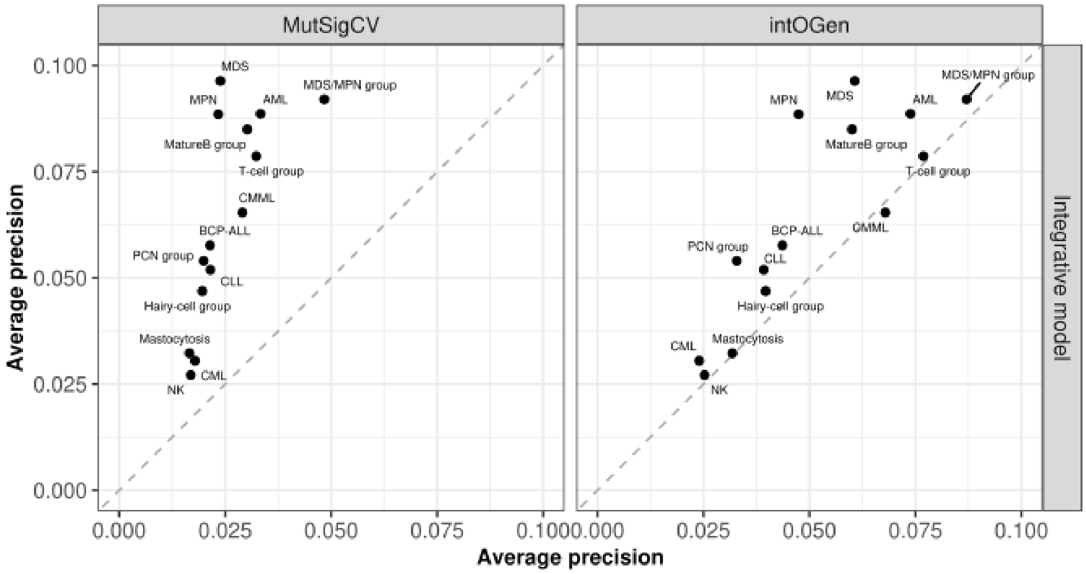


**Supplementary Figure 17. Average precision of the integrative model against MutSigCV and intOGen among study groups.** Across all study groups, our integrative model outperformed or was on par with intOGen or MutSigCV.

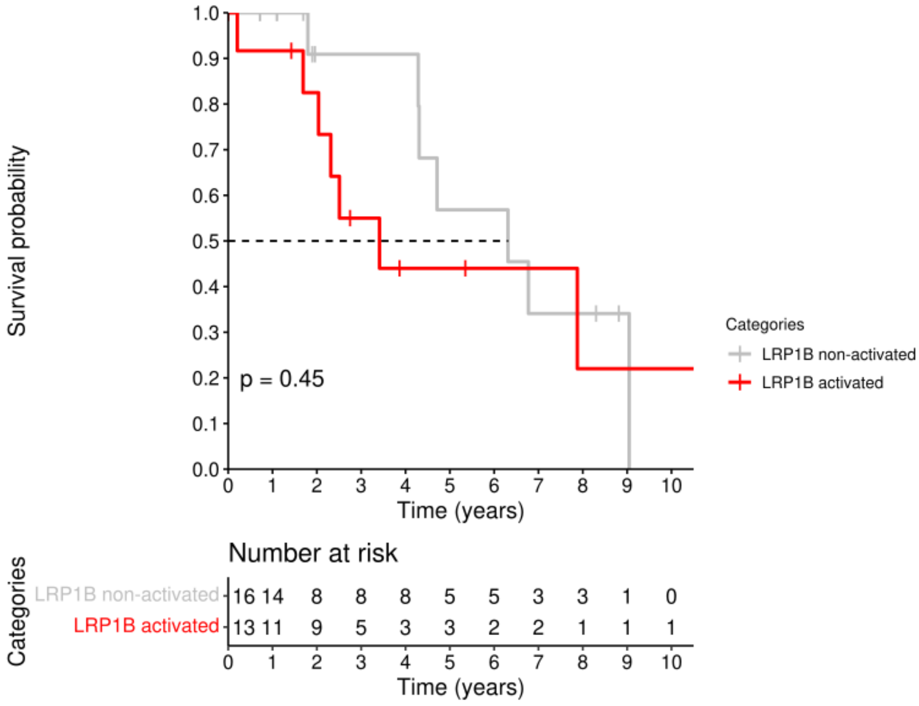


**Supplementary Figure 18. Kaplan-Meier curve estimating survival between *LRP1B*-activated and non-activated samples within HCL-V.** A trend towards shorter overall survival of patients with *LRP1B* activation is observed.

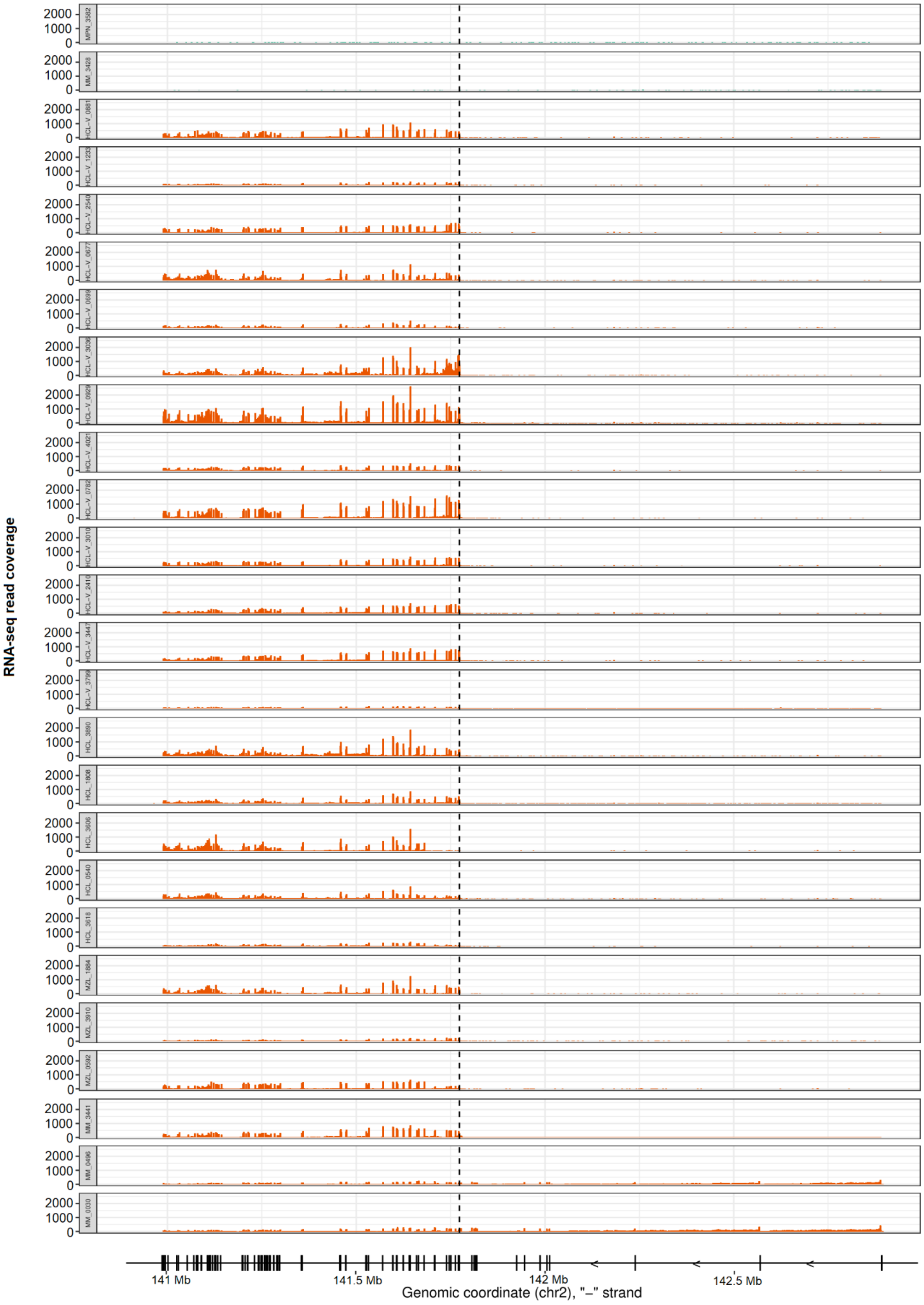


**Supplementary Figure 19. RNA-seq read coverage of *LRP1B* in all *LRP1B*-activated samples (orange) and two *LRP1B*-non-activated examples (green, top 2 rows) in the dataset.** Gene annotation of *LRP1B* (GRCh37) is shown at the bottom.

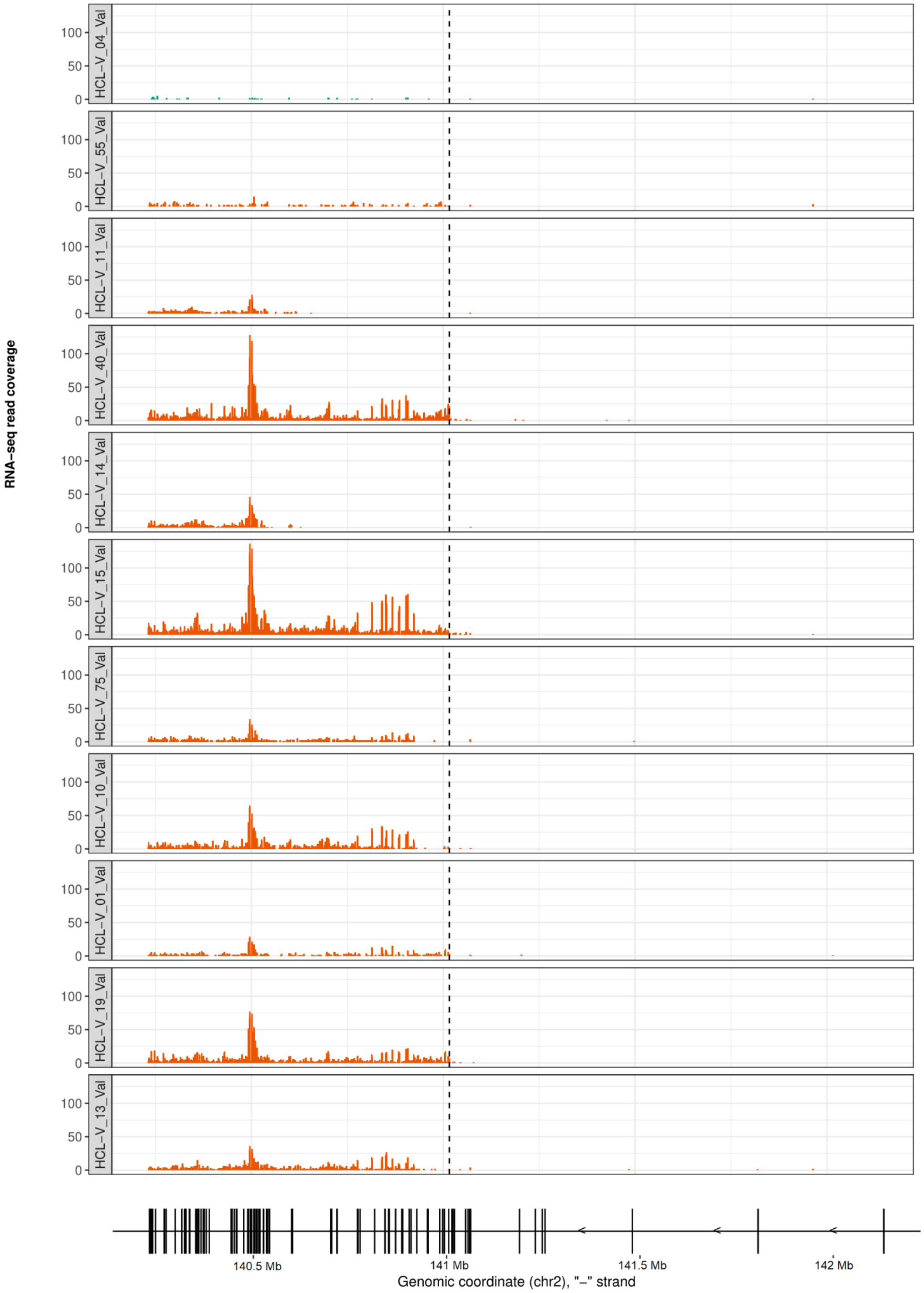


**Supplementary Figure 20. RNA-seq read coverage of *LRP1B* in all *LRP1B*-activated samples (orange) and one *LRP1B*-non-activated example (green, top row) in the validation dataset.** Gene annotation of *LRP1B* (GRCh38) is shown at the bottom.
